# The MIND Study: Design, Feasibility, and Baseline Characteristics of a Smartphone-Based Migraine Cohort

**DOI:** 10.64898/2026.04.14.26350866

**Authors:** Babak Khorsand, Devin Teichrow, Richard B. Lipton, Ali Ezzati

## Abstract

**Objective:** To describe the design, feasibility, and baseline characteristics of the Migraine Impact on Neurocognitive Dynamics (MIND) study, a 30-day smartphone-based cohort for high-frequency assessment of cognition and symptoms in adults with migraine.

**Background:** Cognitive symptoms are an important component of migraine burden, but they are difficult to measure using single-visit testing or retrospective questionnaires. Repeated smartphone-based assessment may better capture real-world variability in cognition and symptoms.

**Methods:** Adults meeting International Classification of Headache Disorders, 3rd edition, criteria for migraine were enrolled remotely and completed 30 days of once-daily ecological momentary assessments and mobile cognitive tasks delivered through the Mobile Monitoring of Cognitive Change platform. Baseline measures assessed demographics, migraine characteristics, disability, mood, stress, and treatment patterns. Feasibility was evaluated using enrollment, completion, and retention metrics.

**Results:** A total of 177 participants enrolled (mean age 38.8 ± 11.9 years; 79.7% female), including 80/177 (45.2%) with chronic migraine. Across the 30-day protocol, 3688 daily assessments were completed, representing 70.8% of all possible study days, and 70.6% of participants completed at least 20 days of monitoring. Completion remained above 60% across study days. At baseline, chronic migraine was associated with greater burden than low-frequency and high-frequency episodic migraine, including higher MIDAS scores (98.6 vs. 38.7 and 70.3), more days with concentration difficulty (16.0 vs. 7.9 and 11.5), and more days with functional interference (18.5 vs. 7.6 and 13.0).

**Conclusions:** The MIND study demonstrates the feasibility of high-frequency smartphone-based assessment of cognition and symptoms in migraine and provides a methodological foundation for future analyses of within-person cognitive and symptom dynamics across the migraine cycle.

## Introduction

Migraine is a common and disabling neurologic disorder whose burden extends beyond headache to include sensory symptoms, fatigue, mood changes, and cognitive difficulties ^1^. Cognitive symptoms, often described as “brain fog,” slowed thinking, trouble concentrating, and forgetfulness, are increasingly recognized as a meaningful component of migraine burden and an important treatment target.^2, 3^ Patients report these symptoms across multiple phases of the migraine cycle, including the premonitory, ictal, postdrome, and interictal periods, and often describe them as disruptive to work, daily functioning, and self-management ^4, 5^.

Despite their clinical relevance, migraine-related cognitive symptoms remain difficult to measure in a way that reflects real-world experience. Two conventional approaches to the assessment of cognition in migraine exist, and each has complementary but important limitations. First, objective cognitive testing is often administered during a single clinic or laboratory visit; while standardized, it captures only a brief snapshot under controlled conditions and may miss the day-to-day and within-day variability that patients report across migraine phases and daily contexts. Second, many studies characterize cognition and related symptoms using self-report questionnaires that ask participants to summarize experiences over weeks to months. These retrospective reports are inherently subjective and vulnerable to recall bias, and they tend to collapse heterogeneous daily experiences into a single “average” estimate, limiting insight into when cognitive difficulties emerge, how they relate to symptom phase, and which contextual factors, such as sleep disruption, stress, mood changes, and medication use, coincide with symptom worsening.

A key methodological implication is that cognition in migraine should not be treated solely as a static trait measured at a single point in time. A growing clinical and neurobiological literature supports the view of migraine as a cyclic condition with phase-dependent changes, including during the premonitory phase and postdrome, and persistent interictal burden in many individuals^6, 7^. For outcomes such as cognition, mood, and daily functioning, in which both subjective experience and objective performance are influenced by fluctuating state factors, dense sampling can help separate stable between-person differences that reflect who tends to report or perform worse overall from within-person dynamics that reflect when a given individual deviates from his or her own typical functioning. The latter is particularly relevant to migraine, where the clinical question is often not only “who is impaired,” but “when, under what conditions, and in which phase does functioning change?”^8^

Smartphone-based repeated assessment offers a practical and scalable path to capturing these dynamics in daily life. Ecological momentary assessment (EMA) can capture symptoms and context close in time to the experience, reducing reliance on retrospective summaries and enabling time-locked analyses around phase transitions^9^. In parallel, brief, repeatable mobile cognitive tasks can provide objective indices of cognitive efficiency that can be sampled frequently with manageable burden^10, 11^.

Pairing these two measurement modes in the same protocol allows the field to move beyond symptom diaries alone. Objective cognitive performance can be interpreted alongside concurrent pain, sensory sensitivity, mood, stress, sleep-related factors, medication use, and functional interference, variables that are central to how patients experience “brain fog” and how clinicians conceptualize disability.

Headache research has increasingly demonstrated the value of decentralized, app-based cohorts for characterizing migraine populations and treatment patterns at scale ^8, 12–14^. However, a standardized, migraine-focused methodological framework that integrates high-frequency symptom and context assessment with repeated objective cognitive measurement remains relatively underdeveloped. A methods-forward cohort report in this space must address design challenges that are specific to high-frequency cognitive measurement in real-world settings, including balancing temporal resolution against participant burden, selecting tasks that are brief and repeatable without excessive learning effects, ensuring interpretability of daily symptom and context measures, and reporting sampling and compliance features transparently to support reproducibility and cross-study comparability. Established reporting guidance for momentary assessment studies emphasizes the importance of clearly describing monitoring period, sampling design, technology, and compliance metrics, principles that become even more important when cognition is a primary repeated outcome.

The Migraine Impact on Neurocognitive Dynamics (MIND) study was designed to address this methodological gap. MIND is a prospective, decentralized, smartphone-based cohort developed to capture daily objective cognitive performance alongside repeated reports of migraine symptoms and relevant contextual factors in real-world settings. Although cognitive testing is a central focus, the study design recognizes that cognition in migraine is shaped by symptom phase and daily context. In this first paper from the MIND study, we (1) describe the study design and measurement strategy, (2) evaluate feasibility and engagement with the 30-day protocol, and (3) summarize baseline cohort characteristics. Together, these findings provide a methodological foundation for future analyses of within-person cognitive and symptom dynamics across the migraine cycle.

## Methods

### Study Design and Setting

The Migraine Impact on Neurocognitive Dynamics (MIND) study is a prospective, decentralized, smartphone-based observational study designed to measure daily cognitive performance in real-world settings alongside momentary migraine symptoms and contextual factors. This report describes the study design and measurement strategy, summarizes baseline characteristics, and evaluates feasibility and engagement with a 30-day repeated-assessment protocol. The study was approved by the University of California, Irvine Institutional Review Board, and all participants provided informed consent prior to participation.

Eligibility criteria were intentionally broad to support recruitment of adults with migraine across a range of symptom burden and backgrounds. Participants were eligible if they were adults aged 18 to 65 years, were English-speaking, had access to a compatible smartphone with reliable internet, and met screening criteria for migraine based on International Classification of Headache Disorders, 3rd edition (ICHD-3), criteria. Key exclusions included major neurologic disorders other than migraine and current alcohol or drug abuse. Full inclusion and exclusion criteria are presented in Supplementary Table 1.

### Recruitment and Enrollment

Participants were recruited nationwide using decentralized digital outreach, including online advertisements and migraine-focused online communities (e.g., migraine-specific Reddit subreddits) between October 2024 and August 2025. Interested individuals completed an online screening form hosted on the study website. A total of 1,074 individuals completed the screening questionnaire. Of these, 421 met eligibility criteria, 186 enrolled, and 177 completed baseline surveys and initiated study participation, forming the final baseline cohort (Figure 1).

**Figure 1.**
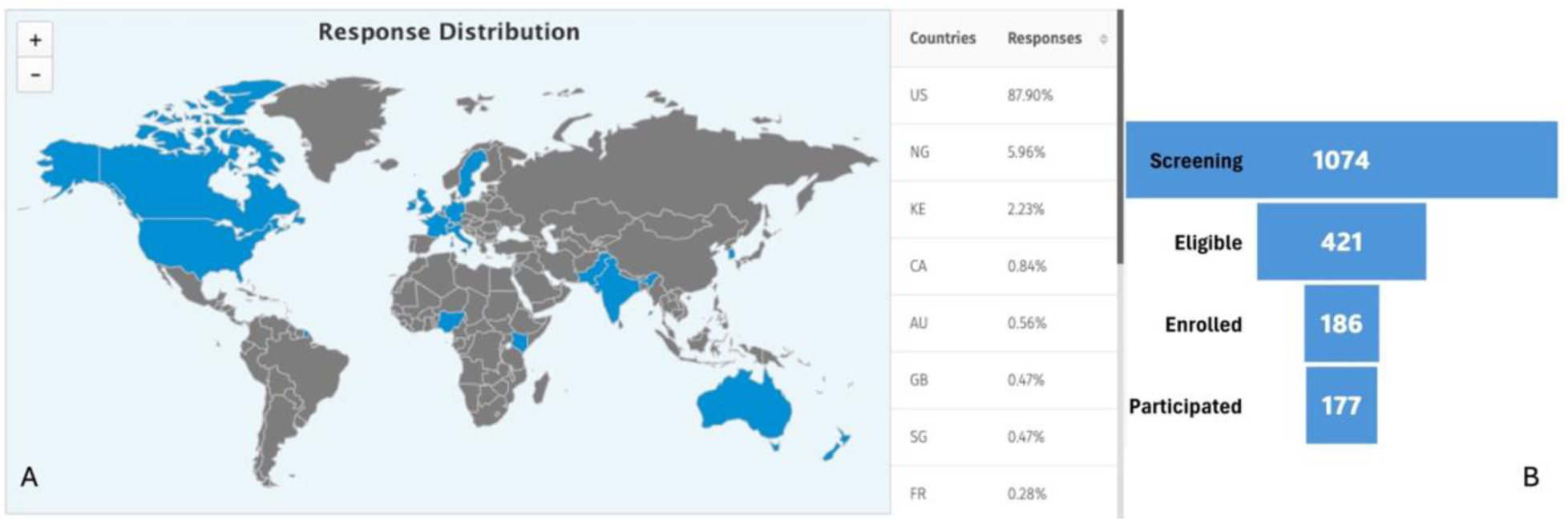
**A)** Choropleth map showing the distribution of users across the world in screening phase. **B)** Study participant flow from screening to final participation.

#### Study procedures and assessment schedule

Participants completed one smartphone-based assessment per day for 30 days using MetricWire, a secure, HIPAA-compliant mobile platform. Daily assessments were available from 12:00 PM to 10:00 PM local time to minimize interference with morning routines and to capture a consistent daily period when symptoms, fatigue, and cognitive functioning commonly affect activities. Participants received a daily prompt at the start of the time period and, if the assessment was not completed, hourly reminder notifications were delivered within the completion window until the daily session was finished or the time window closed. Each daily assessment included migraine symptom and context questions followed by brief cognitive tasks. Participants who remained active were invited to complete an end-of-study questionnaire battery at Day 30. The schedule and measure domains collected at baseline, daily, and Day 30 are summarized in Figure 2.

**Figure 2).**
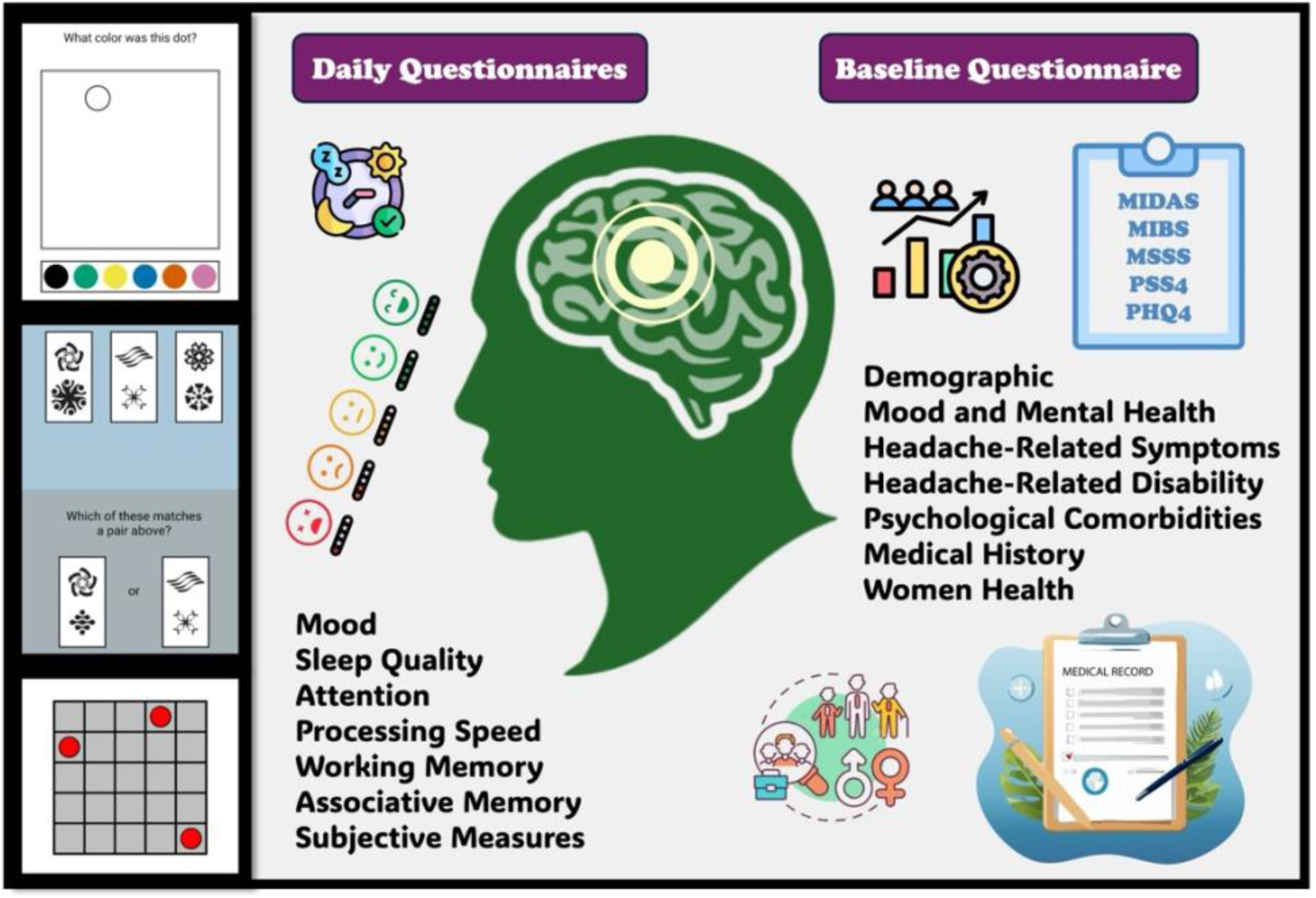
Visualization of the various domains for daily and baseline questionnaires *Note: Baseline and Day 30 questionnaires assessed overlapping domains, while daily assessments focused on momentary symptoms and context*.

### Baseline and end-of-study questionnaires

At enrollment, participants completed questionnaires assessing the following domains:

- Sociodemographics: age, sex, race/ethnicity, education, employment.
- Migraine characteristics: ICHD-3 subtype, attack frequency, typical duration, and pain features
- Disability and burden: Migraine Disability Assessment Scale (MIDAS)^15^, Migraine Severity Scale Score (MSSS)^16^, Migraine Interictal Burden Scale (MIBS) ^17^, and Migraine Treatment Optimization Questionnaire-6 (mTOQ-6)^18^.
- Affective and stress measures: Patient Health Questionnaire-4 (PHQ-4)^19^ and Perceived Stress Scale-4 (PSS-4)^20^.

Instruments were selected for comparability with prior digital migraine studies such as the Headache Assessment via Digital Platform in the United States (HeAD-US) and Observational survey of the Epidemiology, Treatment, and Care of Migraine (OVERCOME) studies^12, 21, 22^. Participants were invited to complete a similar questionnaire battery at Day 30 to characterize the study among those remaining active through the end of the observation period.

#### Daily EMA

Daily items assessed headache status and intensity, mood and irritability, perceived stress, concentration difficulty, medication use, and functional interference. Items were designed to be brief and repeatable to support daily completion over the 30-day monitoring period. The complete day 1, daily, and day 30 item sets and response options are provided in Supplementary Tables 2 and 3.

### Digital Cognitive Tasks

Following the daily surveys, participants completed brief cognitive tasks the Mobile Monitoring of Cognitive Change (M2C2) battery: (1) *Symbol Search* assesses processing speed and visual scanning by requiring participants to determine whether a target symbol is present among comparison symbols. (2) *Color Dots* measures attention and short-term memory through recall of color-location pairings. (3) *Grid Memory* measures visuospatial working memory by asking participants to reproduce briefly presented grid patterns after a short delay. Each task required less than two minutes to complete, allowing the full cognitive component to be completed within a short daily session.

Each task required less than two minutes to complete, allowing the full cognitive component to be completed within a short daily session. These tasks were selected based on prior evidence supporting their feasibility for mobile administration, psychometric performance under repeated testing, and sensitivity to intraindividual variability. To support interpretation of repeated measures, subsequent analyses will explicitly consider practice effects and within-person baselines when modeling cognitive dynamics over the monitoring period^14, 23, 24^.

### Data Management and Quality Control

For this baseline and feasibility report, analyses were based on the full baseline cohort of participants who completed baseline surveys and initiated study participation.

Additional data-quality and analytic-subset criteria were prespecified for subsequent longitudinal cognitive analyses. These included requiring at least 2 headache days and 2 non-headache days during follow-up, excluding trials with reaction times less than 200 milliseconds or greater than 10,000 milliseconds, and excluding participants with mean task accuracy more than 3 standard deviations below the sample mean. These additional criteria were prespecified for subsequent phase-based cognitive analyses and were not used to define the baseline cohort for the present feasibility and baseline report.

### Statistical Analysis

Analyses for this report were primarily descriptive. Continuous variables are summarized as mean (SD) and categorical variables as n (%). When comparing baseline characteristics across migraine-frequency groups (low-frequency episodic, high-frequency episodic, and chronic migraine), continuous outcomes were compared using one-way analysis of variance or a nonparametric alternative when distributional assumptions were not met, and categorical outcomes were compared using chi-square or Fisher’s exact tests as appropriate. When pairwise comparisons were performed, they were conducted using two-sample tests corresponding to the outcome type. All tests were two-sided with alpha set at 0.05 and were considered exploratory; p values were not adjusted for multiple comparisons.

Feasibility and engagement outcomes were summarized descriptively, including day-level completion rates across the 30-day window, number of completed days per participant, the proportion meeting prespecified completion thresholds, and retention through Day 30. Item-level adherence and patterns of missingness are reported in a similar manner. Analyses used complete-case denominators for each variable, and the denominator is reported where it varies. Analyses were performed using R version 4.5.2.

### Data Availability

De-identified data are available from the corresponding author upon reasonable request and subject to institutional data-sharing agreements.

## Results

### Study assembly and baseline cohort

Participants were recruited using decentralized digital outreach from across the United States (Figure 1A). A total of 1,074 individuals completed the screening questionnaire; 421 met eligibility criteria, 186 enrolled, and 177 completed baseline surveys and initiated study participation (Figure 1B). At baseline, 59 participants (33.3%) were classified as low-frequency episodic migraine, 38 (21.5%) as high-frequency episodic migraine, and 80 (45.2%) as chronic migraine.

### Feasibility and engagement with daily assessments

Across the 30-day protocol, 3,688 daily assessments were completed. This corresponds to 70.8% of all possible person-days (3,688 of 5,310 expected daily assessments based on 177 participants over 30 days). Day-level completion remained at or above 60% across study days (Figure 3A).

**Figure 3.**
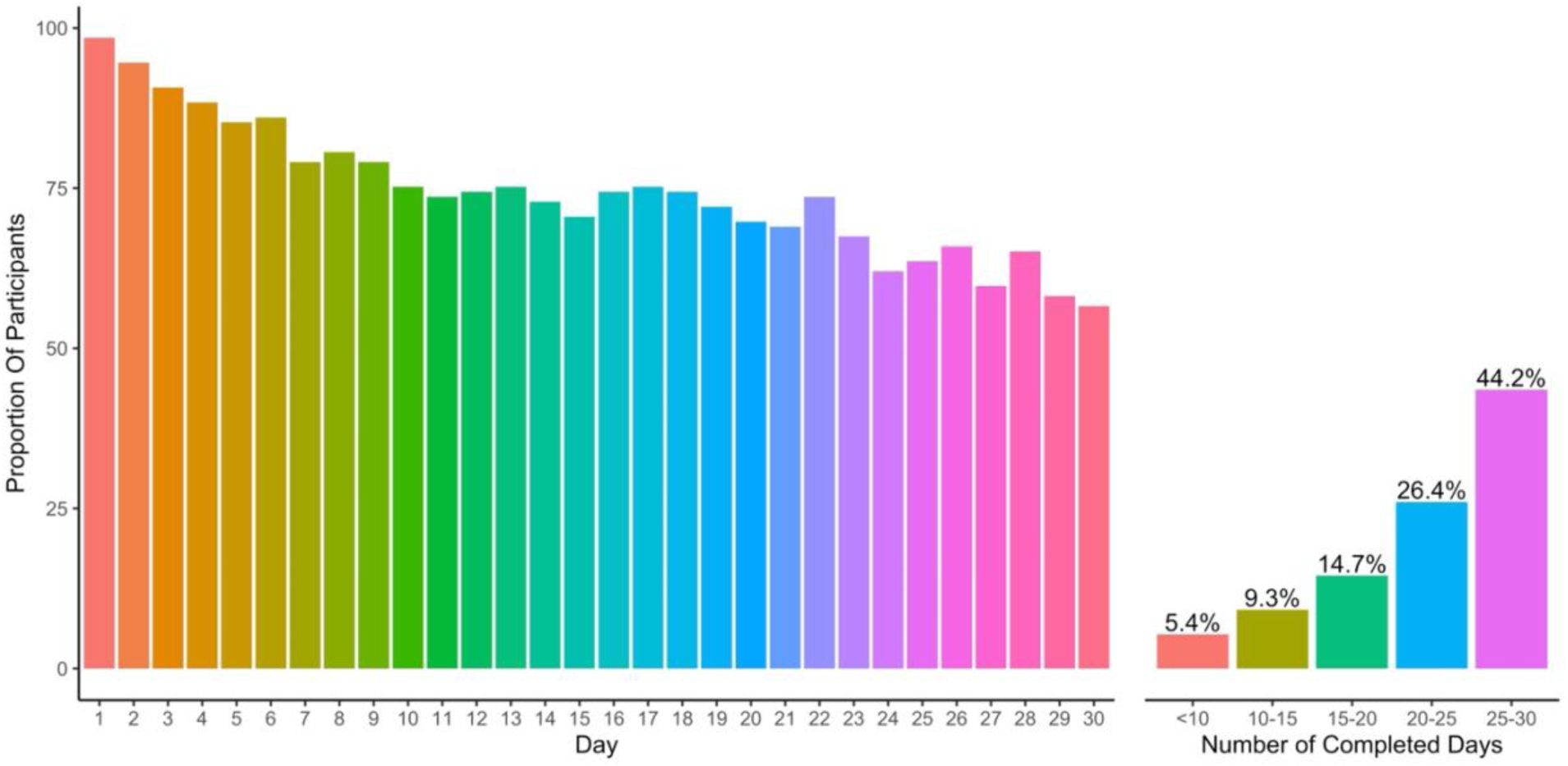
**A)** Proportion of participants who completed each of the 30 days. **B)** Proportion who completed less than 10 days, 10-15 days, 15-20 days, 20-25 days, and 25-30 days; Day-level completion represents the proportion of enrolled participants who completed the full daily assessment on each study day. A completed day was defined as completion of all daily EMA items and all cognitive tasks.

Of the completed surveys, 886 (24.0%) were filled within the first hour after the daily assessment became avaialbe. A total of 1334 (36.2%) were completed within the first two hours, 1988 (53.9%) within the first four hours, and 3019 (81.9%) within the first eight hours (Figure 4).

**Figure 4).**
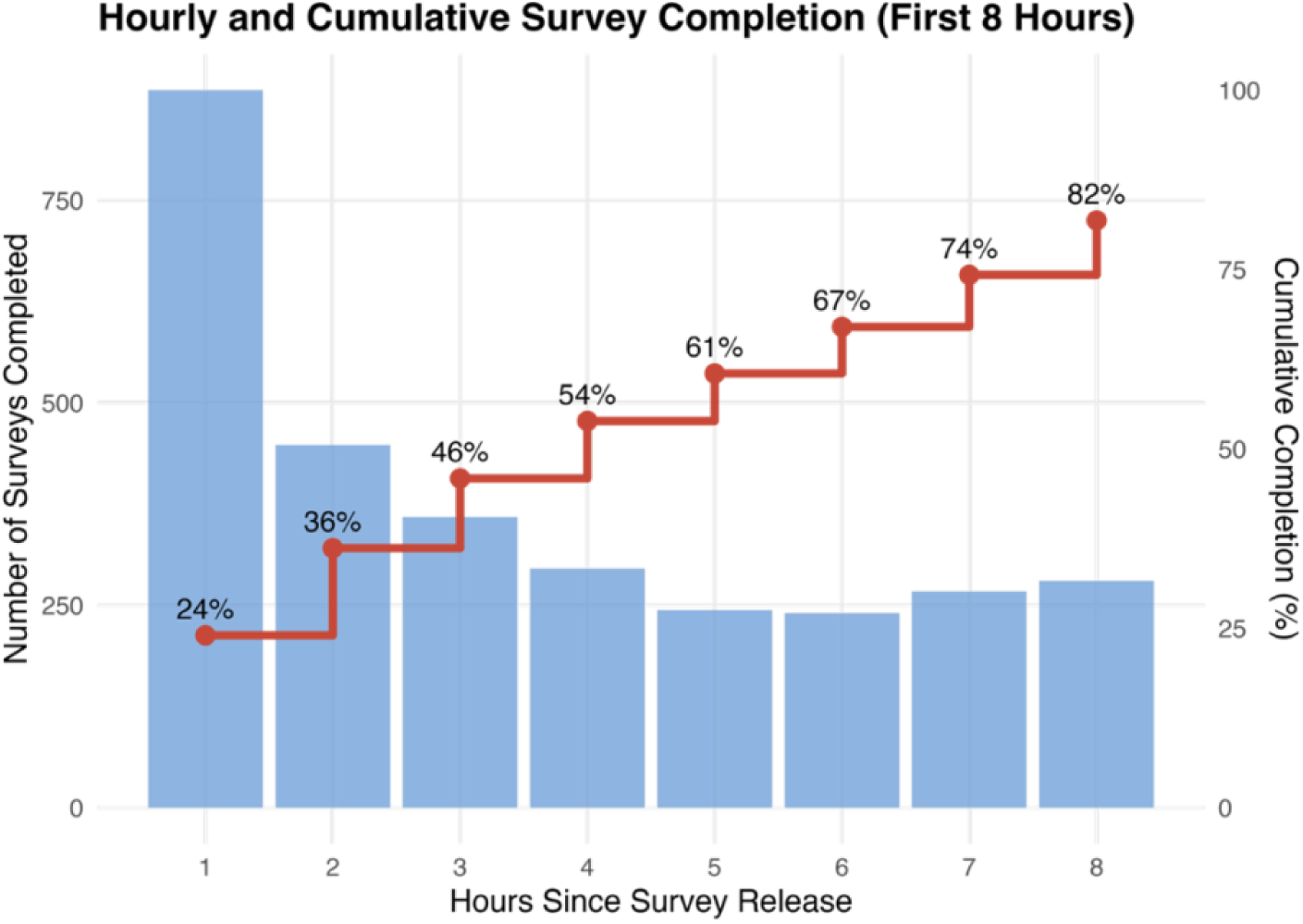
Survey completion behavior during the first eight hours. Blue bars show the number of surveys completed each hour since the survey was released (hourly responses), while the red step curve represents the cumulative percentage of completed surveys. Percentages on the curve indicate cumulative completion at each hour. This figure illustrates both the speed and overall coverage of participant responses. Daily assessments were released at 12:00 PM local time. Hourly bars represent the number of completed surveys within each hour following release; the cumulative curve represents the proportion of completed surveys over time. Hourly reminder notifications were delivered during the completion window if the assessment was not completed.

Correlation analysis between completion rate and the examined variables revealed several modest associations (Figure 5). Among demographic characteristics, completion rates were lower among White participants (r = −0.18, p = 0.03), whereas age, sex, education, marital status, and ethnicity were not significantly associated with completion rate. Among clinical and subjective factors, completion rate was negatively associated with subjective brain fog severity (r = −0.28, p < 0.001) and the proportion of days with headache (r = −0.23, p = 0.004), and positively associated with sleep quality (r = 0.21, p = 0.010). Mood, stress, enjoyment, sleep duration, and pain interference measures were not significantly associated with completion rate (all p > 0.05).

**Figure 5).**
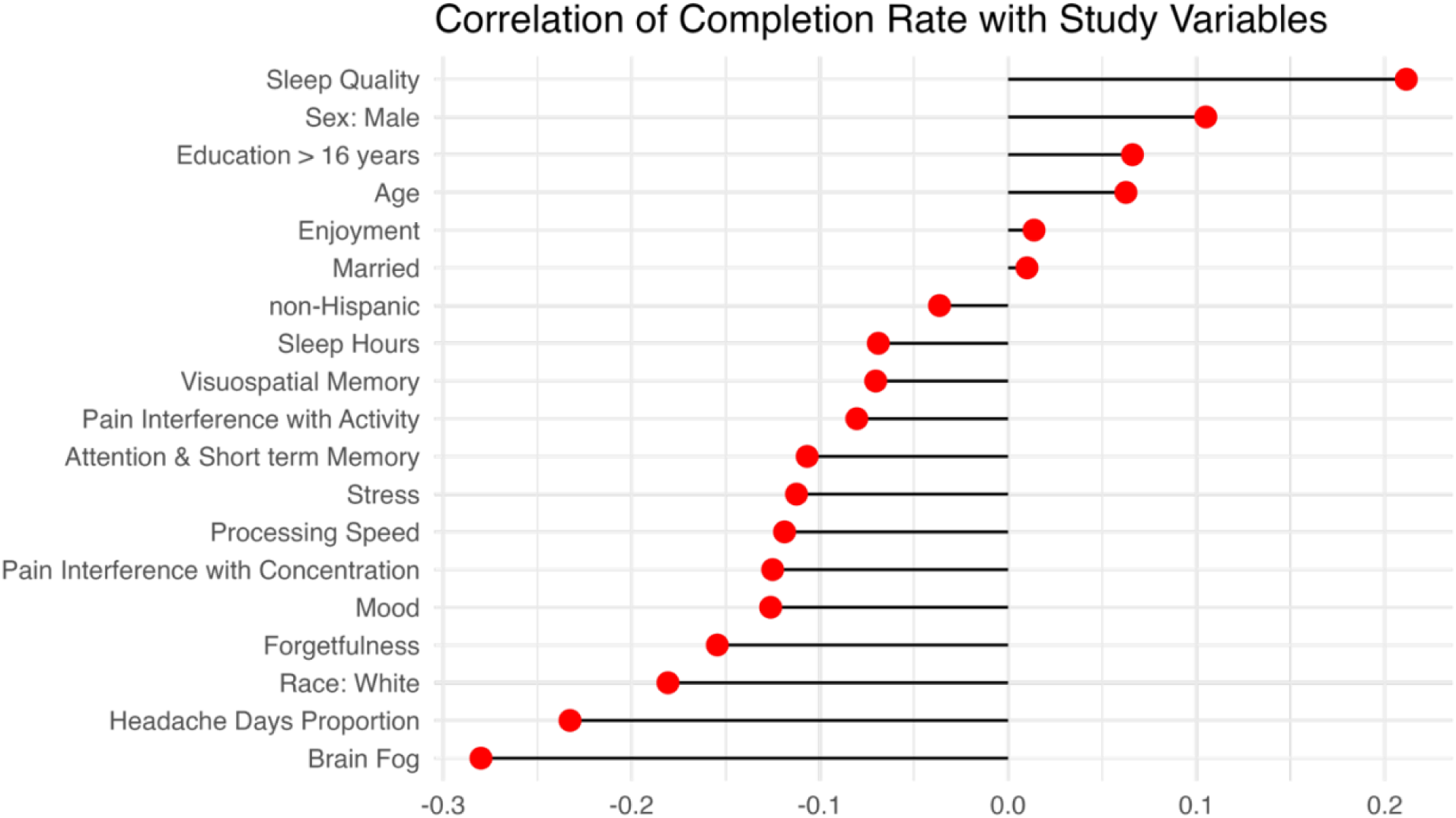
Lollipop plot illustrating the Pearson correlation coefficients between completion rate and the examined study variables. Each point represents the correlation coefficient for a specific variable, while the horizontal line indicates the direction and magnitude of the relationship. Positive values denote a positive association with completion rate, whereas negative values indicate an inverse association. Variables are ordered according to the strength of their correlation. Completion rate was defined as the proportion of available study days completed by each participant over the 30-day monitoring period. Correlations reflect participant-level associations and are presented for descriptive purposes.

Daily engagement patterns further supported the feasibility of the protocol. Supplementary Table 7 presents item-level EMA completeness. Ninety-three percent (27/29) of the items had a total missingness of less than 2%. Only two items had overallmissingness greater than 2%. Only two items had missingness greater than 2%: the item asking “What time did your headache start?” had 5% missing data, and the item asking participants to “list any medications taken that day for a headache or migraine” had 4% missing data.

In Supplementary Table 8, we report the number of missing records in the daily questionnaires for each participant. Out of 177 participants, 7 (4.0%) had missingness greater than 3%. Six participants (3.4%) had missingness between 1% and 3%, and 60 participants (33.9%) had missingness of less than 1%. The remaining 104 participants (58.8%) had no missing data.

#### Baseline sociodemographic characteristics

The baseline cohort included 177 participants with a mean age of 38.8 ± 11.9 years (range 18–69); 141 (79.7%) were female. Table 1 summarizes the demographic characteristics of the study sample stratified by reported gender.

**Table 1).** Sample characteristics stratified by gender.

|  |  | Overall | Female | Male |
| --- | --- | --- | --- | --- |
| **n (%)** |  | 177 (100%) | 141 (79.7%) | 36 (20.3%) |
| **Age, Mean(SD)** |  | 38.84 (11.86) | 39.30 (11.74) | 37.06 (12.32) |
| **Marital Status (%)** | Single | 57 (32.2%) | 42 (29.8%) | 15 (41.7%) |
|  | Living with partner | 21 (11.9%) | 17 (12.1%) | 4 (11.1%) |
|  | Married | 90 (50.8%) | 75 (53.2%) | 15 (41.7%) |
|  | Divorced | 4 (2.3%) | 2 (1.4%) | 2 (5.6%) |
|  | Separated | 2 (1.1%) | 2 (1.4%) | 0 (0.0%) |
|  | Widowed | 1 (0.6%) | 1 (0.7%) | 0 (0.0%) |
|  | Prefer not to answer | 2 (1.1%) | 2 (1.4%) | 0 (0.0%) |
| **Income (%)** | \$25000-50000 | 19 (10.7%) | 9 (6.4%) | 10 (27.8%) |
| | \$50000-75000 | 21 (11.9%) | 13 (9.2%) | 8 (22.2%) |
| | \$75000-100000 | 24 (13.6%) | 20 (14.2%) | 4 (11.1%) |
| | >\$100000 | 75 (42.4%) | 66 (46.8%) | 9 (25.0%) |
|  | Prefer not to answer | 38 (21.5%) | 33 (23.4%) | 5 (13.9%) |
| **Education (%)** | High school or less (<12years) | 3 (1.7%) | 3 (2.1%) | 0 (0.0%) |
|  | High school and less than College (12-16 years) | 25 (14.1%) | 20 (14.2%) | 5 (13.9%) |
|  | College degree or more (>16 years) | 148 (83.6%) | 117 (83.0%) | 31 (86.1%) |
|  | Prefer not to answer | 1 (0.6%) | 1 (0.7%) | 0 (0.0%) |
| **Employment (%)** | Student | 10 (5.6%) | 9 (6.4%) | 1 (2.8%) |
|  | Not employed and not looking for a job | 3 (1.7%) | 3 (2.1%) | 0 (0.0%) |
|  | Long-term or short-term disability | 10 (5.6%) | 10 (7.1%) | 0 (0.0%) |
|  | Homemaker | 10 (5.6%) | 10 (7.1%) | 0 (0.0%) |
|  | Looking for a job | 8 (4.5%) | 7 (5.0%) | 1 (2.8%) |
|  | Employed part time | 28 (15.8%) | 22 (15.6%) | 6 (16.7%) |
|  | Employed full time | 102 (57.6%) | 76 (53.9%) | 26 (72.2%) |
|  | Retired | 4 (2.3%) | 3 (2.1%) | 1 (2.8%) |
|  | Prefer not to answer | 2 (1.1%) | 1 (0.7%) | 1 (2.8%) |
| **Hispanic (%)** | No | 167 (94.4%) | 132 (93.6%) | 35 (97.2%) |
|  | Yes | 8 (4.5%) | 7 (5.0%) | 1 (2.8%) |
|  | Prefer not to answer | 2 (1.1%) | 2 (1.4%) | 0 (0.0%) |
| **Race (%)** | White or Caucasian only | 143 (80.8%) | 121 (85.8%) | 22 (61.1%) |
|  | American Indian or Alaska Native only | 1 (0.6%) | 1 (0.7%) | 0 (0.0%) |
|  | Asian only | 5 (2.8%) | 5 (3.5%) | 0 (0.0%) |
|  | Black or African American Only | 21 (11.9%) | 9 (6.4%) | 12 (33.3%) |
|  | Others-Two or more races | 5 (2.8%) | 3 (2.1%) | 2 (5.6%) |
|  | Prefer not to answer | 2 (1.1%) | 2 (1.4%) | 0 (0.0%) |

#### Baseline migraine characteristics and functional burden

Table 2 summarizes the headache characteristics, functional impact, and psychological features stratified by migraine frequency. CM participants comprised the largest subgroup (n = 80; 45.2%). Across all groups, average sleep duration was roughly seven hours per night and reported pain intensity was consistently high. Sensory features, including allodynia and aura, increased with migraine frequency; nearly half of CM participants reported frequent allodynia versus roughly one in five in the LFEM group.

**Table 2).** Patient reported outcomes stratified by migraine type.

| **Migraine Frequency** |  | **Low Frequency Episodic Migraine** | **High Frequency Episodic Migraine** | **Chronic Migraine** |
| --- | --- | --- | --- | --- |
| **Headache Characteristics** |  |  |  |  |
| Total |  | 59 (33.3%) | 38 (21.5%) | 80 (45.2%) |
| Sleep, Mean (SD) |  | 6.85 (1.06) | 7.19 (1.15) | 6.99 (1.15) |
| Pain, Mean (SD) |  | 8.10 (1.59) | 8.24 (1.77) | 8.55 (1.30) |
| MSSS total score, Mean (SD) |  | 25.12 (2.50) | 24.95 (2.65) | 25.24 (2.79) |
| Allodynia (%) | Half the time or more | 12 (20.3) | 9 (24.3) | 36 (45.0) |
|  | Less than half the time | 19 (32.2) | 15 (40.5) | 20 (25.0) |
|  | Rarely | 18 (30.5) | 11 (29.7) | 13 (16.2) |
|  | Never | 10 (16.9) | 2 (5.4) | 11 (13.8) |
| Aura (%) | Half the time or more | 20 (33.9) | 6 (16.2) | 25 (31.2) |
|  | Less than half the time | 14 (23.7) | 11 (29.7) | 23 (28.7) |
|  | Rarely | 14 (23.7) | 14 (37.8) | 16 (20.0) |
|  | Never | 11 (18.6) | 6 (16.2) | 16 (20.0) |
| Warning before headache (%) | Half the time or more | 27 (46.6) | 16 (43.2) | 44 (55.0) |
|  | Less than half the time | 19 (32.8) | 12 (32.4) | 12 (15.0) |
|  | Rarely | 10 (17.2) | 5 (13.5) | 18 (22.5) |
|  | Never | 2 (3.4) | 4 (10.8) | 6 (7.5) |
| Numbness before headache (%) | Half the time or more | 7 (11.9) | 4 (10.8) | 15 (19.0) |
|  | Less than half the time | 15 (25.4) | 13 (35.1) | 18 (22.8) |
|  | Rarely | 17 (28.8) | 10 (27.0) | 23 (29.1) |
|  | Never | 20 (33.9) | 10 (27.0) | 23 (29.1) |
| **Headache Impact and Disability** |  |  |  |  |
| Days interfered Concentrate, Mean (SD) |  | 7.63 (5.66) | 13.00 (7.16) | 15.97 (8.59) |
| Days interfered Work, Study, Life, Mean (SD) |  | 7.95 (6.21) | 11.51 (6.84) | 18.45 (8.27) |
| MIDAS total score, Mean (SD) |  | 38.71 (27.43) | 70.28 (54.87) | 98.57 (71.87) |
| MIDAS categories | None | 4 (7.0) | 1 (2.8) | 3 (4.0) |
|  | Mild | 2 (3.5) | 0 (0.0) | 4 (5.3) |
|  | Moderate | 7 (12.3) | 1 (2.8) | 0 (0.0) |
|  | Severe | 44 (77.2) | 34 (94.4) | 68 (90.7) |
| MIBS total score, Mean (SD) |  | 14.97 (2.99) | 15.54 (3.86) | 15.93 (4.00) |
| MIBS categories | None | 0 (0.0) | 1 (2.8) | 0 (0.0) |
|  | Mild | 5 (8.9) | 1 (2.8) | 9 (12.0) |
|  | Moderate | 50 (89.3) | 28 (77.8) | 56 (74.7) |
|  | Severe | 1 (1.8) | 6 (16.7) | 10 (13.3) |
| **Psychological and Reproductive Factors** |  |  |  |  |
| PHQ4 total score, Mean (SD) |  | 3.32 (2.53) | 3.86 (2.91) | 4.44 (3.00) |
| PHQ4 categories | None | 23 (39.0) | 15 (40.5) | 21 (26.2) |
|  | Mild | 24 (40.7) | 12 (32.4) | 34 (42.5) |
|  | Moderate | 10 (16.9) | 7 (18.9) | 16 (20.0) |
|  | Severe | 2 (3.4) | 3 (8.1) | 9 (11.2) |
| PSS4 total score, Mean (SD) |  | 12.47 (1.57) | 13.11 (2.01) | 12.60 (1.72) |
| Likelihood Menstrual Headache (%) | Much less likely | 3 (8.8) | 0 (0.0) | 1 (2.3) |
|  | Somewhat less likely | 1 (2.9) | 1 (4.3) | 1 (2.3) |
|  | Same as other times | 13 (38.2) | 8 (34.8) | 17 (38.6) |
|  | Somewhat more likely | 6 (17.6) | 8 (34.8) | 3 (6.8) |
|  | Much more likely | 11 (32.4) | 6 (26.1) | 22 (50.0) |
| Ever pregnant (%) | No | 16 (47.1) | 15 (65.2) | 28 (59.6) |
|  | Yes, in the past 5 years | 4 (11.8) | 5 (21.7) | 5 (10.6) |
|  | Yes, >5 years ago | 14 (41.2) | 3 (13.0) | 14 (29.8) |
| First trimester change (%) | Significantly decreased | 5 (27.8) | 2 (25.0) | 2 (10.5) |
|  | Moderately decreased | 3 (16.7) | 0 (0.0) | 1 (5.3) |
|  | No change | 6 (33.3) | 2 (25.0) | 12 (63.2) |
|  | Moderately increased | 1 (5.6) | 1 (12.5) | 2 (10.5) |
|  | Significantly increased | 3 (16.7) | 3 (37.5) | 2 (10.5) |

*Note: MSSS = Migraine Symptom Severity Score; MIDAS = Migraine Disability Assessment Score; MIBS = Migraine Interictal Burden Scale; PHQ4 = Patient Health Questionnaire-4; PSS4 = Perceived Stress Scale-4; Migraine frequency groups were defined as low-frequency episodic migraine (2–7 migraine days/month), high-frequency episodic migraine (8–14 migraine days/month), and chronic migraine ( $\geq 15$ migraine days/month). Values represent baseline self-reported measures unless otherwise indicated. Reproductive history items were calculated among participants reporting menstrual history.*

Warning symptoms also followed this pattern.

Participants with chronic migraine exhibited greater disability and impairment across multiple domains. MIDAS scores differed significantly among the three groups, with the CM group reporting the highest mean total score (98.56), exceeding both the HFEM group (70.28, p = 0.02) and the LFEM group (38.71, p < 0.001); all three group means were within the severe disability range. HFEM participants also had significantly higher MIDAS scores than LFEM participants (p = 0.002). Psychological symptoms showed a similar pattern: PHQ-4 scores were higher in the CM group (4.44) than in the LFEM group (3.32, p = 0.02). Measures of functional interference also showed a strong gradient by migraine frequency. The CM group reported more days with concentration difficulties (15.97) than both the HFEM group (11.51, p < 0.001) and the LFEM group (7.94, p < 0.001), and HFEM participants exceeded LFEM participants on the same measure (p = 0.01). Days with interference in work, study, or daily life also differed significantly, with the CM group reporting the greatest impact (18.45), followed by the HFEM (13.00, p = 0.05) and LFEM (7.62, p < 0.001) groups. Consistent with these patterns, aura frequency varied significantly across groups based on chi-square tests (overall p = 0.01), with CM participants more frequently endorsing aura symptoms than LFEM participants (p = 0.01).

Among participants reporting menstrual history, menstrual-related worsening of migraine was common, particularly in CM, where approximately half endorsed substantially increased likelihood of headaches around menses. Reproductive histories were broadly consistent with the expected demographics for a digitally recruited adult migraine sample.

#### Baseline treatment patterns

Because subgroup sizes were not selected to support formal hypothesis testing for medication differences, these results are presented to characterize treatment patterns and support the clinical face validity of the sample. Preventive treatment complexity generally increased with migraine frequency, particularly in chronic migraine. Use of onabotulinumtoxinA was 18.6% in low-frequency episodic migraine, 23.7% in high-frequency episodic migraine, and 32.5% in chronic migraine; corresponding values for preventive gepants were 20.3%, 18.4%, and 30.0%. Other traditional preventive classes, including beta blockers, antidepressants, and antiseizure medications, were also common and were generally used more often in chronic migraine than in low-frequency episodic migraine (Table 3). Prior preventive medication exposure showed similar gradients and suggested more extensive prior preventive treatment in chronic migraine (Supplementary Table 4).

**Table 3).** Patterns of preventive medication use among participants stratified by episodic (low and high frequency) and chronic migraine.

| Medication | Low Frequency Episodic Migraine | High Frequency Episodic Migraine | Chronic Migraine |
| --- | --- | --- | --- |
| Total | 59 (33.3%) | 38 (21.5%) | 80 (45.2%) |
| Anti-CGRP | 25 (42.4%) | 25 (65.8%) | 34 (42.5%) |
| Gepants | 12 (20.3%) | 7 (18.4%) | 24 (30%) |
| Botox | 11 (18.6%) | 9 (23.7%) | 26 (32.5%) |
| Anti-seizure | 6 (10.2%) | 6 (15.8%) | 9 (11.2%) |
| Anti-depressants | 8 (13.6%) | 3 (7.9%) | 16 (20%) |
| Beta blockers | 4 (6.8%) | 4 (10.5%) | 14 (17.5%) |
| Other medications | 3 (5.1%) | 3 (7.9%) | 5 (6.2%) |

*Note: Low Frequency Episodic Migraine = 2-7 days with migraine per month; High Frequency Episodic Migraine = 8-14 days with migraine per month; Chronic Migraine = 15+ days with migraine per month; CGRP = Calcitonin Gene-Related Peptide; Preventive medication use reflects current self-reported use at baseline. Participants could report more than one preventive medication class; therefore, percentages may sum to greater than 100%.*

Triptans were used by 40.7% of participants with low-frequency episodic migraine, 28.9% with high-frequency episodic migraine, and 48.8% with chronic migraine, while corresponding values for acute gepants were 42.4%, 39.5%, and 47.5%. Over-the-counter analgesic use was also frequent at 33.9%, 50.0%, and 46.2%, respectively. Opioid- or barbiturate-containing medications were uncommon overall but were reported more often in chronic migraine at 3.4%, 0%, and 10.0%, respectively, whereas ergot derivatives were rarely used. Acute monotherapy was used by a minority of participants, suggesting that combination acute treatment was common, particularly in higher-frequency groups. Neuromodulation device use was infrequent overall but was concentrated in chronic migraine at 6.8%, 2.6%, and 16.3%, respectively, with supraorbital transcutaneous electrical nerve stimulation the most commonly reported device at 3.4%, 2.6%, and 10.0%, respectively (Table 4). Prior acute medication exposure broadly mirrored current use, with high lifetime exposure to over-the-counter agents and triptans and greater exposure to several classes in chronic migraine; prior opioid/barbiturate exposure was more common than current use (Supplementary Table 5).

**Table 4).** Patterns of acute medication use among participants stratified by episodic (low and high frequency) and chronic migraine.

| Medication | Low Frequency Episodic Migraine | High Frequency Episodic Migraine | Chronic Migraine |
| --- | --- | --- | --- |
| Total | 59 (33.3%) | 38 (21.5%) | 80 (45.2%) |
| **Acute medication use monotherapy or polytherapy** |  |  |  |
| OTCs | 20 (33.9%) | 19 (50%) | 37 (46.2%) |
| Gepants | 25 (42.4%) | 15 (39.5%) | 38 (47.5%) |
| Triptans | 24 (40.7%) | 11 (28.9%) | 39 (48.8%) |
| Opioids or Barbiturates | 2 (3.4%) | 0 (0%) | 8 (10%) |
| Ergot derivative | 3 (5.1%) | 0 (0%) | 3 (3.8%) |
| Other medications | 4 (6.8%) | 5 (13.2%) | 10 (12.5%) |
| **Acute medication use monotherapy** |  |  |  |
| OTCs | 2 (3.4%) | 3 (7.9%) | 2 (2.5%) |
| Gepants | 9 (15.3%) | 4 (10.5%) | 8 (10%) |
| Triptans | 10 (16.9%) | 3 (7.9%) | 8 (10%) |
| Opioids or Barbiturates | 0 (0%) | 0 (0%) | 1 (1.2%) |
| Ergot derivative | 1 (1.7%) | 0 (0%) | 0 (0%) |
| Other medications | 2 (3.4%) | 0 (0%) | 1 (1.2%) |
| **Medical device use** |  |  |  |
| Any medical device | 4 (6.8%) | 1 (2.6%) | 13 (16.3%) |
| Supraorbital TENS | 2 (3.4%) | 1 (2.6%) | 8 (10%) |
| External vagal nerve stimulation | 0 (0%) | 0 (0%) | 1 (1.2%) |
| Armband remote electrical neuromodulation | 2 (3.4%) | 0 (0%) | 2 (2.5%) |
| TMS single pulse transcutaneous magnetic stimulation | 0 (0%) | 0 (0%) | 2 (2.5%) |

*Note: Low Frequency Episodic Migraine = 2-7 days with migraine per month; High Frequency Episodic Migraine = 8-14 days with migraine per month; Chronic Migraine = 15+ days with migraine per month; OTC = Over-the-counter; TENS = Transcutaneous electrical nerve stimulation; TMS = Transcranial magnetic Stimulation; Acute medication use reflects current self-reported use at baseline. Participants could report use of multiple acute medication classes (polytherapy). Monotherapy refers to participants reporting use of a single acute medication class.*

#### Day 30 migraine characteristics and functional burden

Among the 143 participants (81%) who remained active through Day 30, the distribution of migraine-frequency groups was similar to baseline, with chronic migraine comprising the largest subgroup (44%). Headache characteristics, disability, and psychological measures at Day 30 were broadly consistent with baseline patterns, and gradients by migraine frequency persisted (Supplementary Table 6).

## Discussion

The baseline findings from the MIND study demonstrate the feasibility of using remote, smartphone-based assessment protocols to collect daily symptom and cognitive data in real-world settings. Across the 30-day protocol, participants completed 3688 daily assessments, representing 70.8% of all possible study days; 70.6% completed at least 20 days of monitoring, and day-level completion remained above 60% across study days. A decentralized design successfully integrated ecological momentary assessment (EMA) with brief, repeatable cognitive tasks and maintained engagement across a month-long monitoring period. Participants represented a broad range of migraine severity and demographic backgrounds and were similar in age, sex distribution, and disability burden to other digitally recruited migraine cohorts^12, 21, 22^. High levels of engagement confirm that once-daily mobile assessments paired with brief cognitive testing can be implemented in routine life, consistent with prior work demonstrating reliable adherence to daily EMA protocols^13, 25^. Together, these features establish a scalable framework for capturing day-to-day symptom and cognitive experiences in adults with migraine.

Beyond feasibility alone, the 30-day MIND protocol provides a practical demonstration of a reusable methodological framework for high-frequency digital assessment in migraine. Consistent assessment timing, short task duration, and a fixed task sequence supported sustained engagement without exceeding participant burden, while harmonized daily symptom items integrated smoothly with validated M2C2 cognitive tasks within a single measurement structure. Transparent, rule-based quality-control procedures enabled systematic handling of both survey and cognitive data, supporting reproducibility and transferability to other repeated-measures designs. Together, these features constitute a scalable template for capturing within-person symptom and cognitive dynamics in migraine and other fluctuating neurological conditions.

The MIND study also provides a strong foundation for subsequent longitudinal analyses. The combination of daily symptom reports and repeatable cognitive tasks offers an opportunity to investigate temporal relationships among pain, mood, stress, and cognitive performance, all of which are areas that are difficult to study using retrospective or clinic-based designs. As prior work suggests, cognitive symptoms in migraine may fluctuate substantially across the ictal and interictal periods^4, 26^. High-frequency, ecologically valid assessment is therefore likely to improve understanding of these dynamics and may help clarify the circumstances under which cognitive difficulties are most pronounced. mportantly, completion timing clustered within the first several hours of the daily assessment window, suggesting that predictable availability and consistent timing can effectively anchor participation.

Patterns of engagement also provide insight beyond simple feasibility. Completion rate was lower among participants reporting greater brain fog and a higher proportion of headache days, and higher among those reporting better sleep quality, suggesting that missingness may be at least partly state-dependent. These findings have direct implications for both study design and analysis, underscoring the importance of modeling informative missingness rather than treating non-completion as a purely logistical issue. From a protocol optimization perspective, the results suggest that brief daily sessions, fixed assessment windows, and decentralized recruitment strategies can sustain engagement even among individuals with substantial symptom burden, while highlighting opportunities to further tailor timing, prompting, or adaptive sampling in future iterations ^24, 27, 28^.

Several methodological considerations emerge from the design. First, repeated administration of brief cognitive tasks inevitably introduces practice effects, but the MIND study illustrates that these can be addressed analytically rather than treated as a barrier. Modeling strategies that incorporate within-person baselines and time-on-study adjustments allow learning-related improvements to be separated from clinically meaningful day-to-day variability. Second, the choice of a single afternoon–evening assessment window proved feasible and contributed to predictable engagement, yet it also limits the ability to evaluate diurnal fluctuations or morning-specific cognitive changes. Future protocols may consider adaptive or phase-triggered sampling to broaden temporal coverage. Third, the study highlights the value of transparent, rule-based quality-control procedures for mobile cognitive data. Automated checks for implausible response times, inconsistent completion patterns, and timestamp validity create a reproducible analytic foundation and support downstream sensitivity analyses in which stricter QC thresholds or alternative “complete-day” definitions can be applied to assess robustness. Finally, the protocol underscores several considerations intrinsic to EMA research, including the potential for state-dependent missingness, the constrained temporal scope of once-daily self-report, and the possibility of reactivity to repeated symptom monitoring. These features must be explicitly modeled in longitudinal analyses, particularly when evaluating symptom–cognition coupling or pre-ictal changes. Together, these methodological considerations outline a flexible, generalizable blueprint for integrating brief objective cognitive testing into high-frequency digital symptom research ^14, 27, 29, 30^.

The primary construct of interest in this framework is objective cognitive variability and efficiency over time. Brief, repeatable mobile tasks allow cognition to be measured as a dynamic signal that can be examined in relation to concurrent pain, mood, stress, sleep, medication use, and functional interference. This shifts the scientific question from whether individuals with migraine are cognitively impaired on average to when, under what conditions, and in which phases cognitive performance deviates from an individual’s own baseline; questions that are difficult or impossible to address using conventional clinic-based designs ^14^.

Clinical and psychological patterns observed at baseline aligned with established relationships between migraine frequency, disability, and comorbid symptoms.

Participants with chronic migraine had higher disability scores, more days with concentration difficulty, and more days with functional interference than participants with episodic migraine, and treatment patterns reflected greater clinical complexity, including higher use of onabotulinumtoxinA, gepants, and neuromodulation devices. Headache characteristics and disability measures at Day 30 were broadly consistent with baseline patterns among participants who remained active in the study, supporting the stability of the cohort over the observation period. This consistency suggests that the baseline profile reflects enduring features of migraine burden in this sample rather than transient enrollment effects ^31, 32^.

Several limitations should be acknowledged. First, participants were recruited through online platforms and social media communities, which may introduce selection bias toward individuals who are more digitally engaged or more motivated to track their symptoms. The sample may therefore differ from clinic-based or population-based migraine cohorts. Second, all migraine diagnoses were based on self-report rather than clinician-confirmed ICHD-3 evaluation, although the questionnaire items were designed to map onto diagnostic criteria. Third, adherence, while high overall, varied across individuals, and missing data may influence future longitudinal analyses. In digital symptom studies, missingness may also be state-dependent, for example, participants may be less likely to complete assessments during more severe headache days, raising the possibility of informative missingness that will require careful evaluation in longitudinal models. Fourth, because this report focuses solely on baseline characteristics, no causal or temporal inferences can be made from the observed associations. EMA methods also carry inherent limitations, including reliance on self-report obtained within a constrained time window, potential reactivity to repeated symptom monitoring, and reduced ecological coverage during hours outside the daily assessment window. Finally, while the cognitive tasks used in this study have established reliability and sensitivity to within-person variability, performance may still be influenced by environmental factors inherent to remote testing.

Future analyses will use the longitudinal structure of the MIND dataset to examine within-person relationships among pain, mood, stress, sensory sensitivity, and cognitive function. High-frequency data may help clarify the circumstances under which cognitive symptoms are most prominent and how they evolve across the migraine cycle.

Additionally, repeated measures will allow investigation of potential triggers or moderators of symptom variability, as well as assessment of treatment effects or behavioral adaptations over time. As digital health tools become more widely integrated into headache research, cohorts such as MIND will contribute to establishing standards for remote symptom monitoring and digital cognitive assessment in migraine. The study’s second wave of data collection is currently underway. The second wave will include one month of monitoring with twice-daily EMA. This next wave is designed to capture within-day variability, evaluate modality-specific sensitivity to migraine phase, and test whether multimodal digital signals improve the detection or prediction of cognitive changes relative to once-daily sampling.

In summary, the MIND study demonstrates that remote, smartphone-based EMA and cognitive testing are feasible for adults with migraine and yield clinically meaningful baseline data consistent with prior research. These findings provide a robust platform for subsequent longitudinal analyses aimed at improving understanding of day-to-day cognitive and affective experiences in migraine.

## Data Availability Statement

The datasets used and analyzed during the current study are available from the corresponding author upon reasonable request.

---

**Supplementary Table 1).** Inclusion and exclusion criteria for the MIND Study.

| Phase |  |  | n |
| --- | --- | --- | --- |
| Screening |  |  | 1074 |
| Eligible | Inclusion | Age 18 to 65 years | 421 |
|  |  | Ownership of a compatible smartphone |  |
|  |  | English proficiency sufficient to complete app-based surveys and tasks |  |
|  |  | ICHD-3 migraine diagnosis confirmed at screening based on AMPP criteria |  |
|  |  | Self-reported headache frequency of 3 to 22 headache days per month at screening |  |
|  | Exclusion | Neurological disorders other than migraine |  |
|  |  | History of traumatic brain injury |  |
|  |  | Severe psychiatric illness that would interfere with participation |  |
|  |  | Current alcohol or drug abuse |  |
| Enrolled |  | Eligible participants who answered our calls and accepted our invitation to start the study | 186 |
| Participated |  | Enrolled participants who activated their MetricWire code and started the study | 177 |
| Cognitive | Headache | Participants who had at least two headache days and two non-headache days during the study. | 148 |
|  | Outliers | Exclude participants whose cognitive measures are more than 3 standard deviations above or below the mean | 137 |

**Supplementary Table 2.**
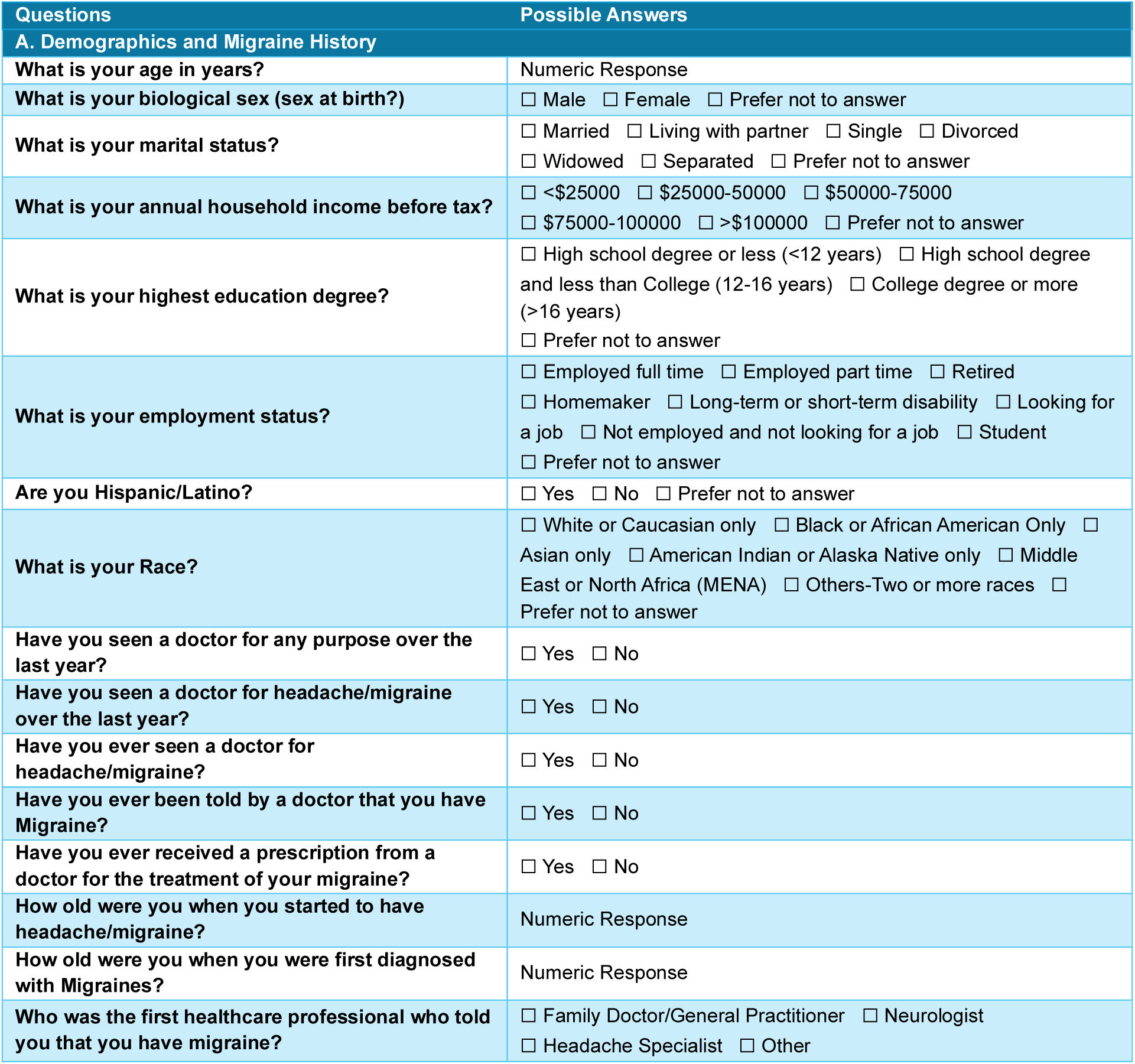
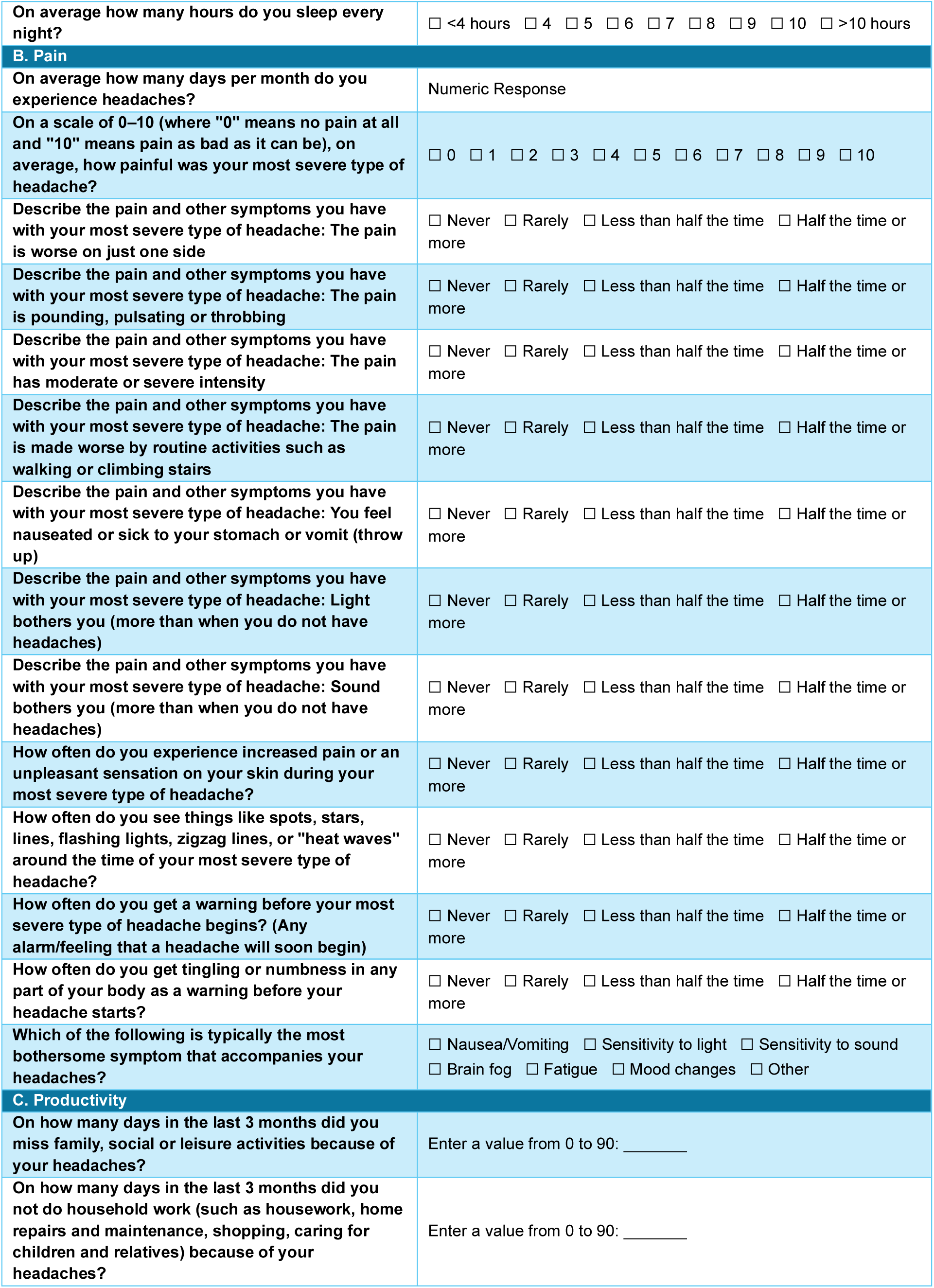
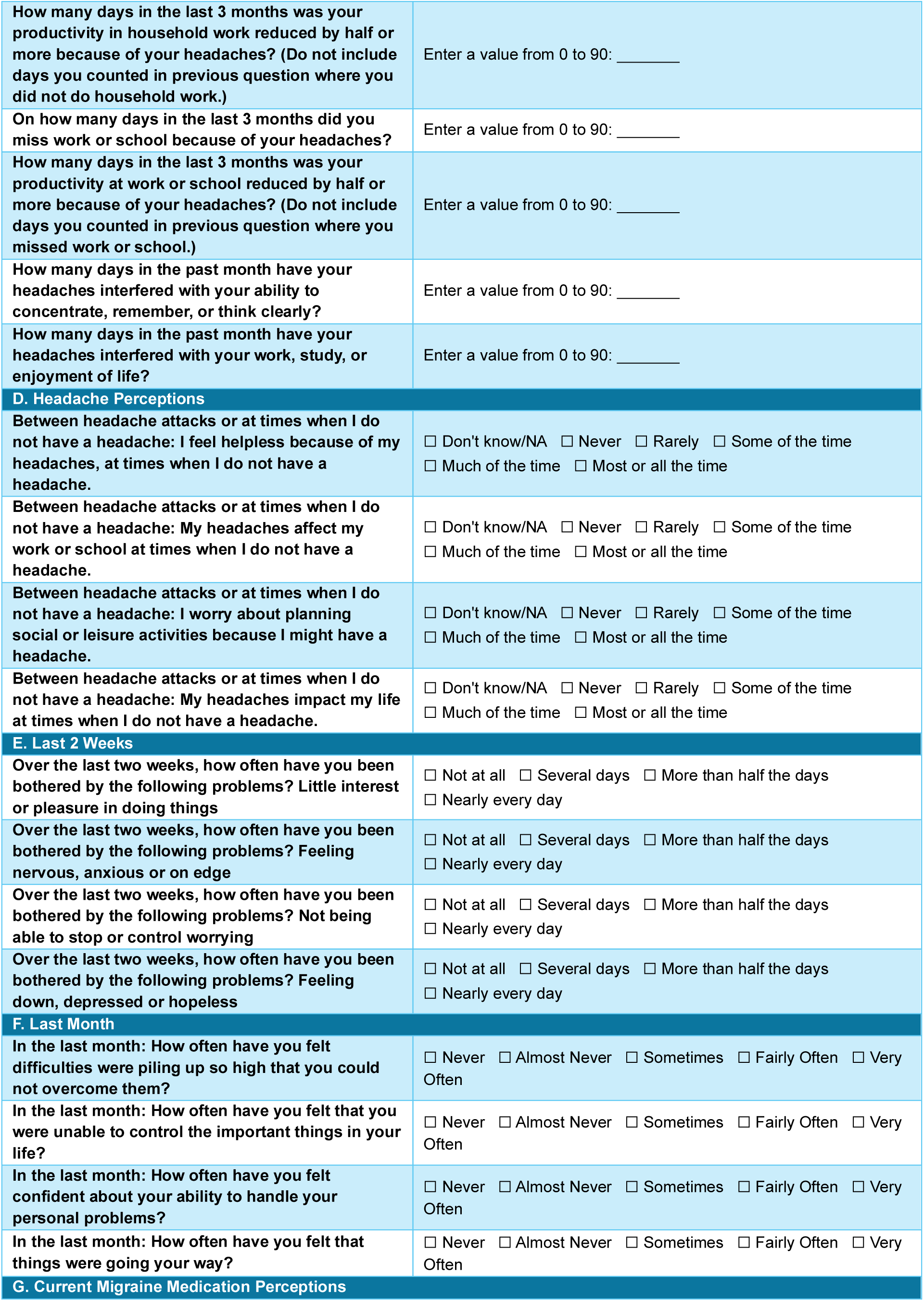
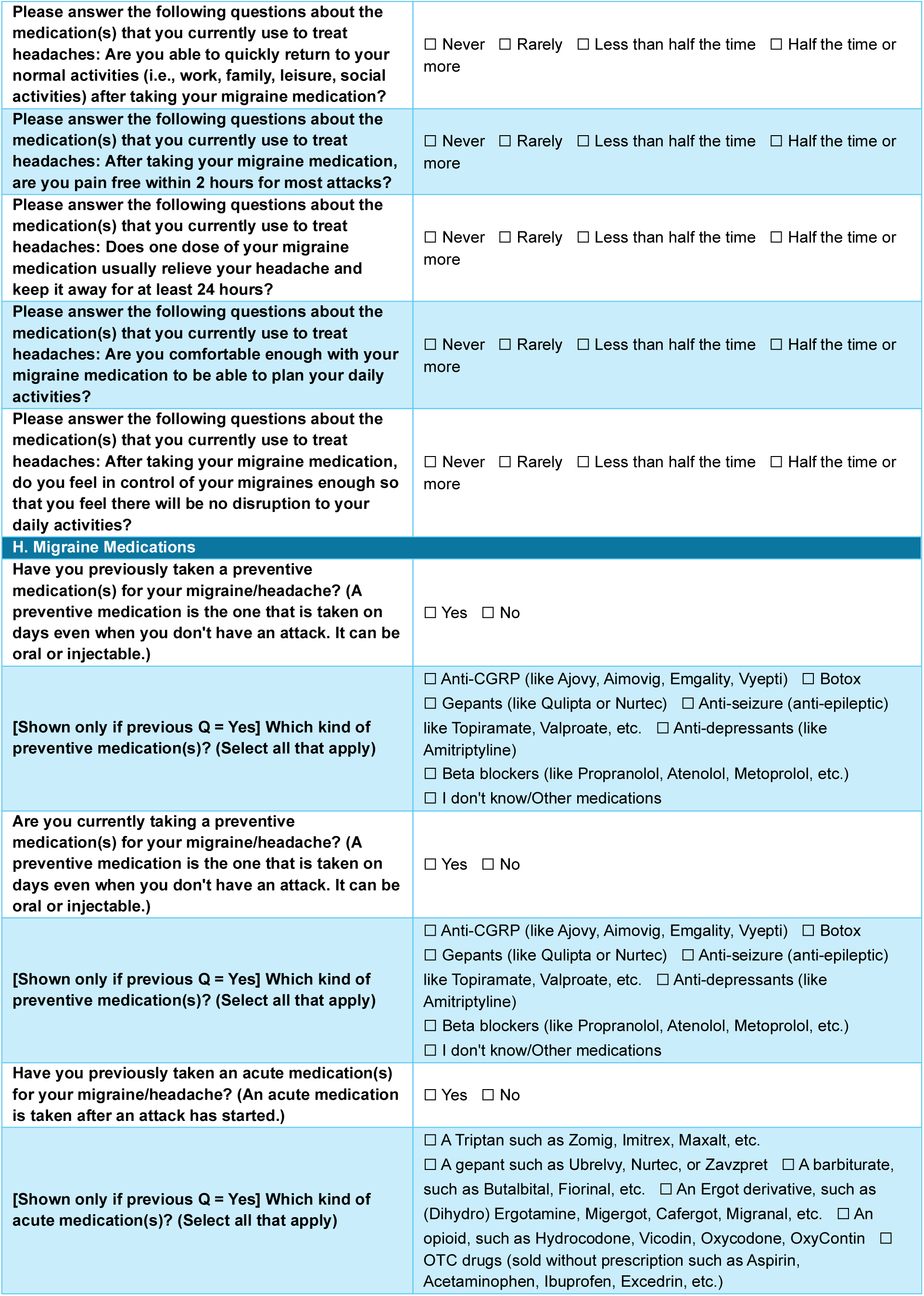
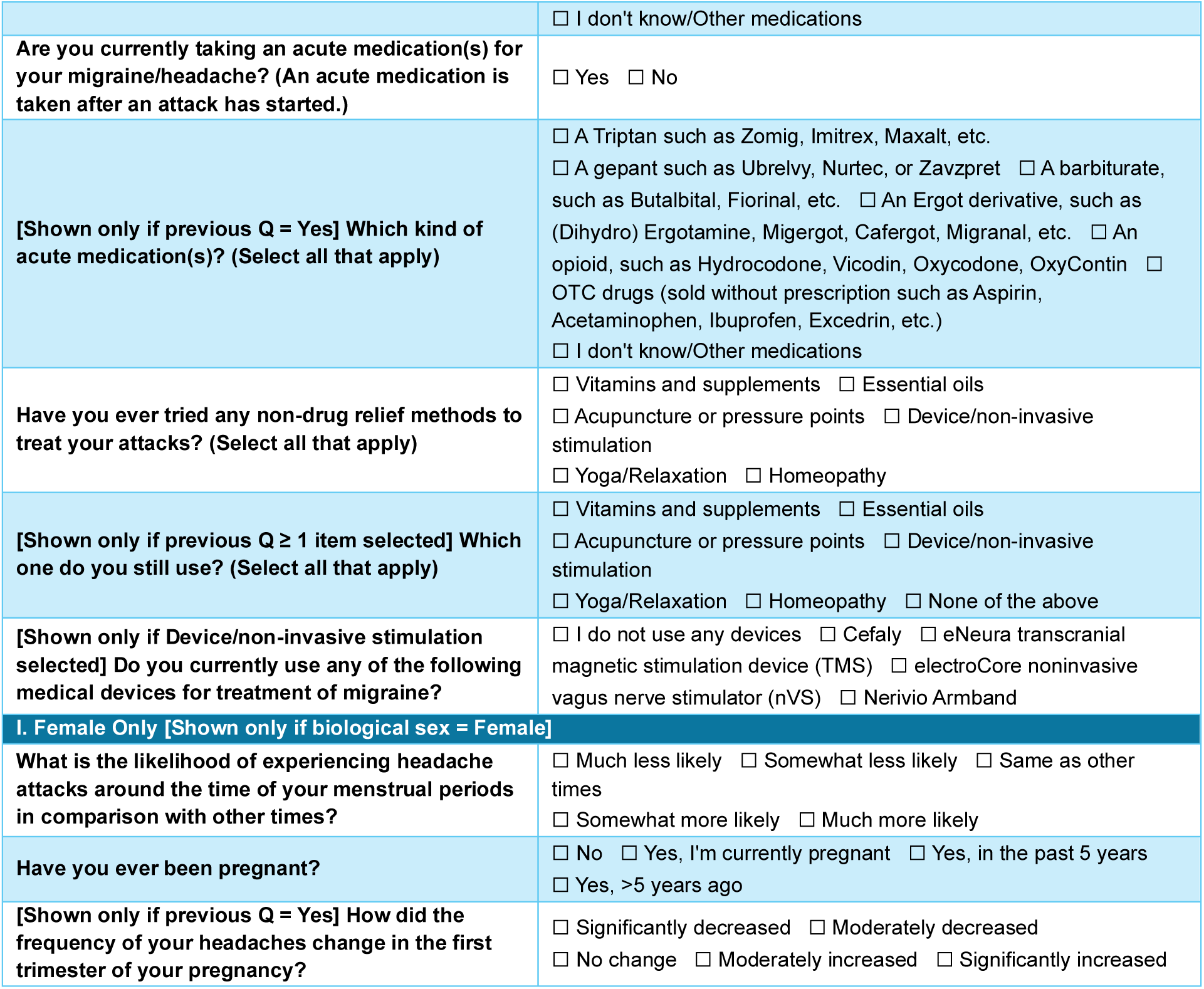
Questionnaire items and response options administered at Day 1. The table provides the full list of survey questions across multiple domains, including demographic, migraine history, headache characteristics, productivity, headache-related perceptions, mood and anxiety symptoms, perceived stress, medication use and effectiveness, and non-pharmacologic treatments. Both categorical and numeric response formats are presented as implemented in the study instrument.

**Supplementary Table 3.**
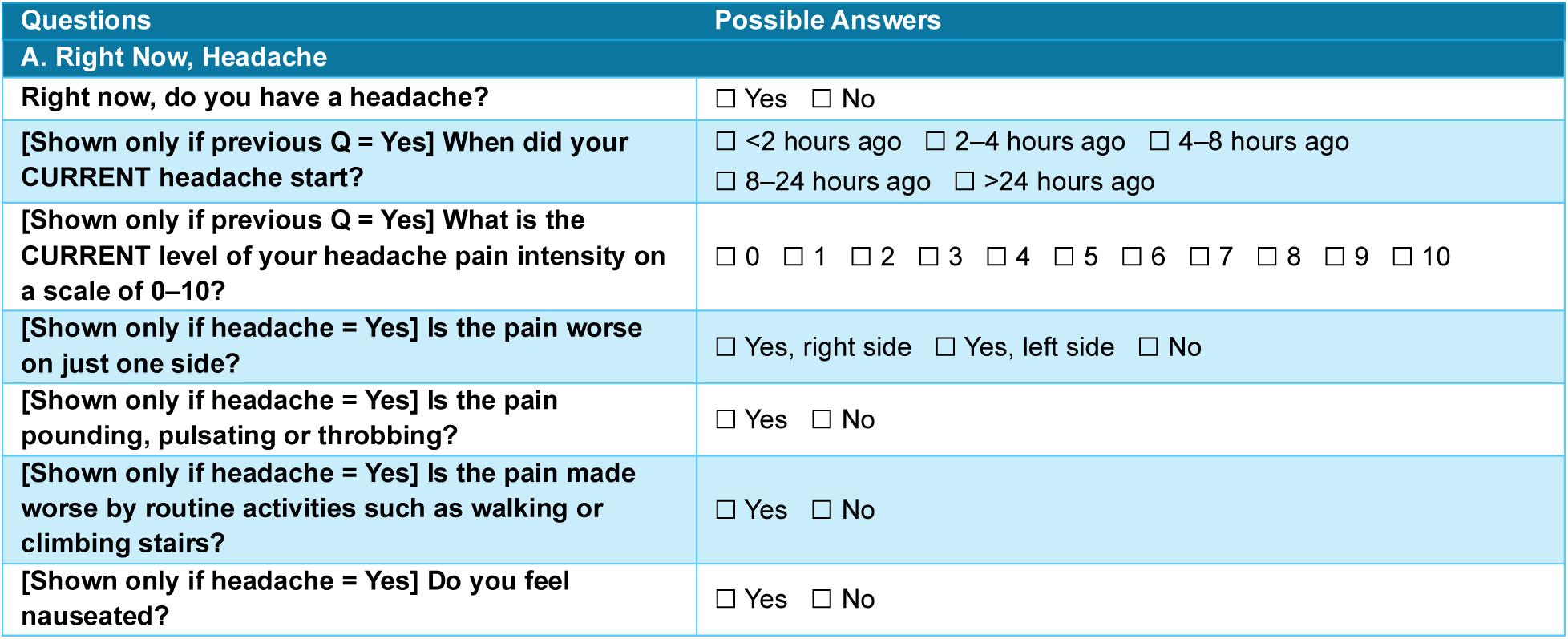
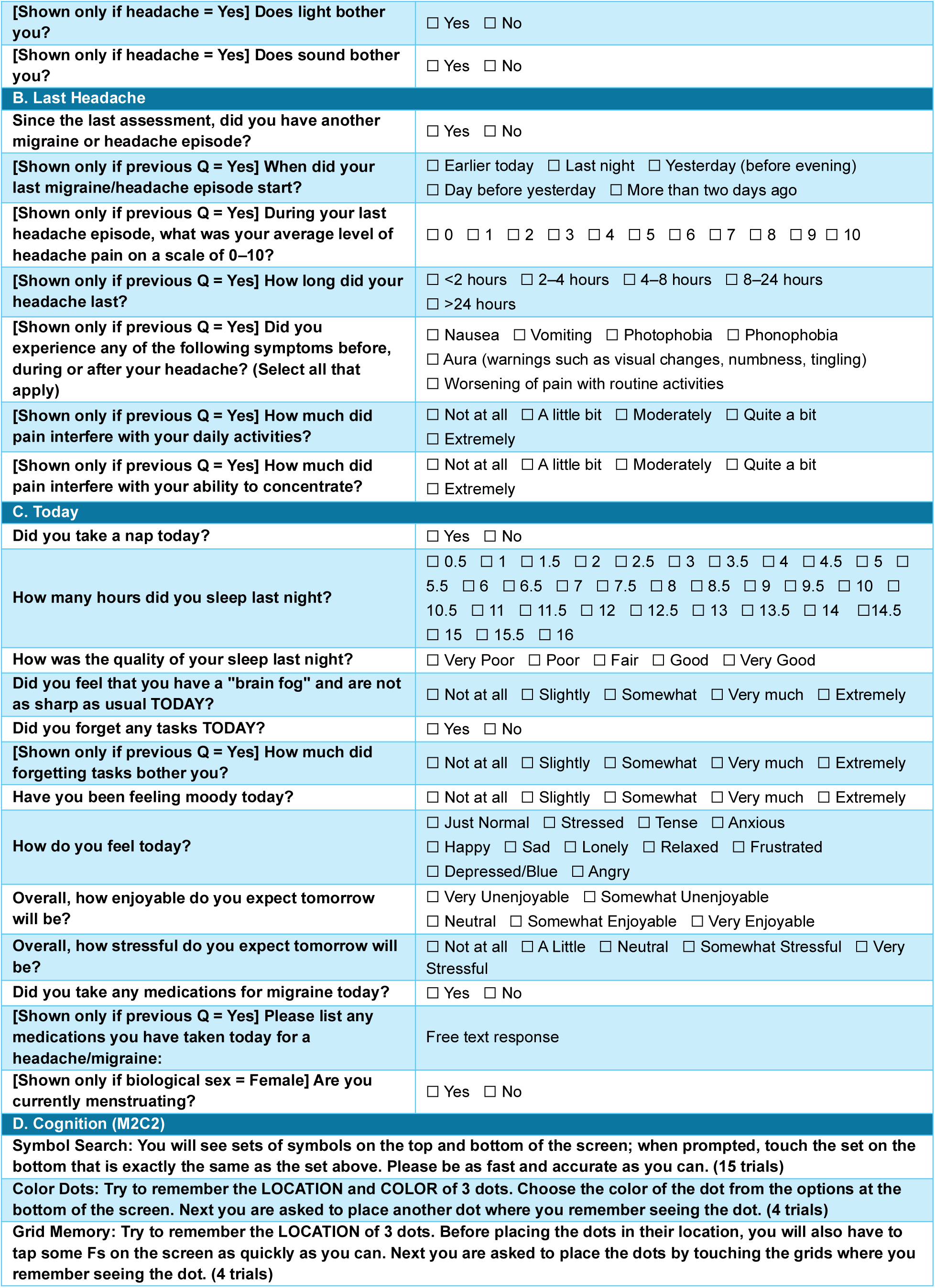
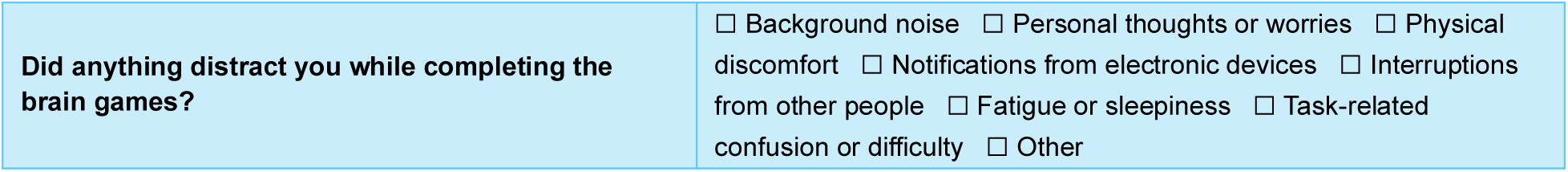
Daily questionnaire items and response options administered once per day over the 30-day study period. The table includes items assessing current headache status, characteristics of the most recent headache episode, daily behaviors and symptoms (e.g., sleep, mood, cognition, and medication use), and performance on mobile cognitive tasks (M2C2). Conditional branching logic was used to display follow-up questions based on participant responses (e.g., presence of headache or medication use).

| Questions | Possible Answers |
| --- | --- |
| **A. Right Now, Headache** |  |
| **Right now, do you have a headache?** | ☐ Yes ☐ No |
| **[Shown only if previous Q = Yes] When did your CURRENT headache start?** | ☐ <2 hours ago ☐ 2–4 hours ago ☐ 4–8 hours ago<br>☐ 8–24 hours ago ☐ >24 hours ago |
| **[Shown only if previous Q = Yes] What is the CURRENT level of your headache pain intensity on a scale of 0–10?** | ☐ 0 ☐ 1 ☐ 2 ☐ 3 ☐ 4 ☐ 5 ☐ 6 ☐ 7 ☐ 8 ☐ 9 ☐ 10 |
| **[Shown only if headache = Yes] Is the pain worse on just one side?** | ☐ Yes, right side ☐ Yes, left side ☐ No |
| **[Shown only if headache = Yes] Is the pain pounding, pulsating or throbbing?** | ☐ Yes ☐ No |
| **[Shown only if headache = Yes] Is the pain made worse by routine activities such as walking or climbing stairs?** | ☐ Yes ☐ No |
| **[Shown only if headache = Yes] Do you feel nauseated?** | ☐ Yes ☐ No |
| [Shown only if headache = Yes] Does light bother you? | ☐ Yes ☐ No |
| [Shown only if headache = Yes] Does sound bother you? | ☐ Yes ☐ No |
| **B. Last Headache** |  |
| Since the last assessment, did you have another migraine or headache episode? | ☐ Yes ☐ No |
| [Shown only if previous Q = Yes] When did your last migraine/headache episode start? | ☐ Earlier today ☐ Last night ☐ Yesterday (before evening)<br>☐ Day before yesterday ☐ More than two days ago |
| [Shown only if previous Q = Yes] During your last headache episode, what was your average level of headache pain on a scale of 0–10? | ☐ 0 ☐ 1 ☐ 2 ☐ 3 ☐ 4 ☐ 5 ☐ 6 ☐ 7 ☐ 8 ☐ 9 ☐ 10 |
| [Shown only if previous Q = Yes] How long did your headache last? | ☐ <2 hours ☐ 2–4 hours ☐ 4–8 hours ☐ 8–24 hours<br>☐ >24 hours |
| [Shown only if previous Q = Yes] Did you experience any of the following symptoms before, during or after your headache? (Select all that apply) | ☐ Nausea ☐ Vomiting ☐ Photophobia ☐ Phonophobia<br>☐ Aura (warnings such as visual changes, numbness, tingling)<br>☐ Worsening of pain with routine activities |
| [Shown only if previous Q = Yes] How much did pain interfere with your daily activities? | ☐ Not at all ☐ A little bit ☐ Moderately ☐ Quite a bit<br>☐ Extremely |
| [Shown only if previous Q = Yes] How much did pain interfere with your ability to concentrate? | ☐ Not at all ☐ A little bit ☐ Moderately ☐ Quite a bit<br>☐ Extremely |
| **C. Today** |  |
| Did you take a nap today? | ☐ Yes ☐ No |
| How many hours did you sleep last night? | ☐ 0.5 ☐ 1 ☐ 1.5 ☐ 2 ☐ 2.5 ☐ 3 ☐ 3.5 ☐ 4 ☐ 4.5 ☐ 5 ☐ 5.5 ☐ 6 ☐ 6.5 ☐ 7 ☐ 7.5 ☐ 8 ☐ 8.5 ☐ 9 ☐ 9.5 ☐ 10 ☐ 10.5 ☐ 11 ☐ 11.5 ☐ 12 ☐ 12.5 ☐ 13 ☐ 13.5 ☐ 14 ☐ 14.5 ☐ 15 ☐ 15.5 ☐ 16 |
| How was the quality of your sleep last night? | ☐ Very Poor ☐ Poor ☐ Fair ☐ Good ☐ Very Good |
| Did you feel that you have a "brain fog" and are not as sharp as usual TODAY? | ☐ Not at all ☐ Slightly ☐ Somewhat ☐ Very much ☐ Extremely |
| Did you forget any tasks TODAY? | ☐ Yes ☐ No |
| [Shown only if previous Q = Yes] How much did forgetting tasks bother you? | ☐ Not at all ☐ Slightly ☐ Somewhat ☐ Very much ☐ Extremely |
| Have you been feeling moody today? | ☐ Not at all ☐ Slightly ☐ Somewhat ☐ Very much ☐ Extremely |
| How do you feel today? | ☐ Just Normal ☐ Stressed ☐ Tense ☐ Anxious<br>☐ Happy ☐ Sad ☐ Lonely ☐ Relaxed ☐ Frustrated<br>☐ Depressed/Blue ☐ Angry |
| Overall, how enjoyable do you expect tomorrow will be? | ☐ Very Unenjoyable ☐ Somewhat Unenjoyable<br>☐ Neutral ☐ Somewhat Enjoyable ☐ Very Enjoyable |
| Overall, how stressful do you expect tomorrow will be? | ☐ Not at all ☐ A Little ☐ Neutral ☐ Somewhat Stressful ☐ Very Stressful |
| Did you take any medications for migraine today? | ☐ Yes ☐ No |
| [Shown only if previous Q = Yes] Please list any medications you have taken today for a headache/migraine: | Free text response |
| [Shown only if biological sex = Female] Are you currently menstruating? | ☐ Yes ☐ No |
| **D. Cognition (M2C2)** |  |
| Symbol Search: You will see sets of symbols on the top and bottom of the screen; when prompted, touch the set on the bottom that is exactly the same as the set above. Please be as fast and accurate as you can. (15 trials) |  |
| Color Dots: Try to remember the LOCATION and COLOR of 3 dots. Choose the color of the dot from the options at the bottom of the screen. Next you are asked to place another dot where you remember seeing the dot. (4 trials) |  |
| Grid Memory: Try to remember the LOCATION of 3 dots. Before placing the dots in their location, you will also have to tap some Fs on the screen as quickly as you can. Next you are asked to place the dots by touching the grids where you remember seeing the dot. (4 trials) |  |
| **Did anything distract you while completing the brain games?** | ☐ Background noise ☐ Personal thoughts or worries ☐ Physical discomfort ☐ Notifications from electronic devices ☐ Interruptions from other people ☐ Fatigue or sleepiness ☐ Task-related confusion or difficulty ☐ Other |

**Supplementary Table 4).** Patterns of previous preventive medication use among participants stratified by episodic (low frequency and high frequency) and chronic migraine.

| Medication | Low Frequency Episodic Migraine | High Frequency Episodic Migraine | Chronic Migraine |
| --- | --- | --- | --- |
| Total | 59 (33.3%) | 38 (21.5%) | 80 (45.2%) |
| Anti-CGRP | 31 (52.5%) | 20 (52.6%) | 58 (72.5%) |
| Gepants | 15 (25.4%) | 14 (36.8%) | 36 (45%) |
| Botox | 14 (23.7%) | 12 (31.6%) | 40 (50%) |
| Anti-seizure | 27 (45.8%) | 20 (52.6%) | 45 (56.2%) |
| Anti-depressants | 21 (35.6%) | 19 (50%) | 51 (63.7%) |
| Beta blockers | 24 (40.7%) | 14 (36.8%) | 43 (53.8%) |
| Other medications | 5 (8.5%) | 3 (7.9%) | 10 (12.5%) |

*Note: Low Frequency Episodic Migraine = 2-7 days with migraine per month; High Frequency Episodic Migraine = 8-14 days with migraine per month; Chronic Migraine = 15+ days with migraine per month; CGRP = Calcitonin Gene-Related Peptide; Previous medication use reflects lifetime self-reported exposure and is not mutually exclusive across medication classes.*

**Supplementary Table 5).** Patterns of previous acute medication use among participants stratified by episodic (low frequency and high frequency) and chronic migraine.

| Medication | Low Frequency Episodic Migraine | High Frequency Episodic Migraine | Chronic Migraine |
| --- | --- | --- | --- |
| Total | 59 (33.3%) | 38 (21.5%) | 80 (45.2%) |
| **Acute medication use monotherapy or polytherapy** |  |  |  |
| OTCs | 38 (64.4%) | 28 (73.7%) | 58 (72.5%) |
| Gepants | 29 (49.2%) | 18 (47.4%) | 48 (60%) |
| Triptans | 42 (71.2%) | 26 (68.4%) | 67 (83.8%) |
| Opioids or Barbiturates | 19 (32.2%) | 18 (47.4%) | 29 (36.3%) |
| Ergot derivative | 8 (13.6%) | 3 (7.9%) | 9 (11.2%) |
| Other medications | 5 (8.5%) | 6 (15.8%) | 7 (8.8%) |
| **Acute medication use monotherapy** |  |  |  |
| OTCs | 0 (0%) | 2 (5.3%) | 1 (1.2%) |
| Gepants | 3 (5.1%) | 0 (0%) | 0 (0%) |
| Triptans | 6 (10.2%) | 2 (5.3%) | 5 (6.2%) |
| Opioids or Barbiturates | 0 (0%) | 1 (2.6%) | 2 (2.5%) |
| Ergot derivative | 1 (1.7%) | 0 (0%) | 0 (0%) |
| Other medications | 0 (0%) | 2 (5.3%) | 0 (0%) |

*Note: Low Frequency Episodic Migraine = 2-7 days with migraine per month; High Frequency Episodic Migraine = 8-14 days with migraine per month; Chronic Migraine = 15+ days with migraine per month; OTC = Over-the-counter; TENS = Transcutaneous electrical nerve stimulation; TMS = Transcranial magnetic Stimulation; Previous medication use reflects lifetime self-reported exposure and is not mutually exclusive across medication classes.*

**Supplementary Table 6).**
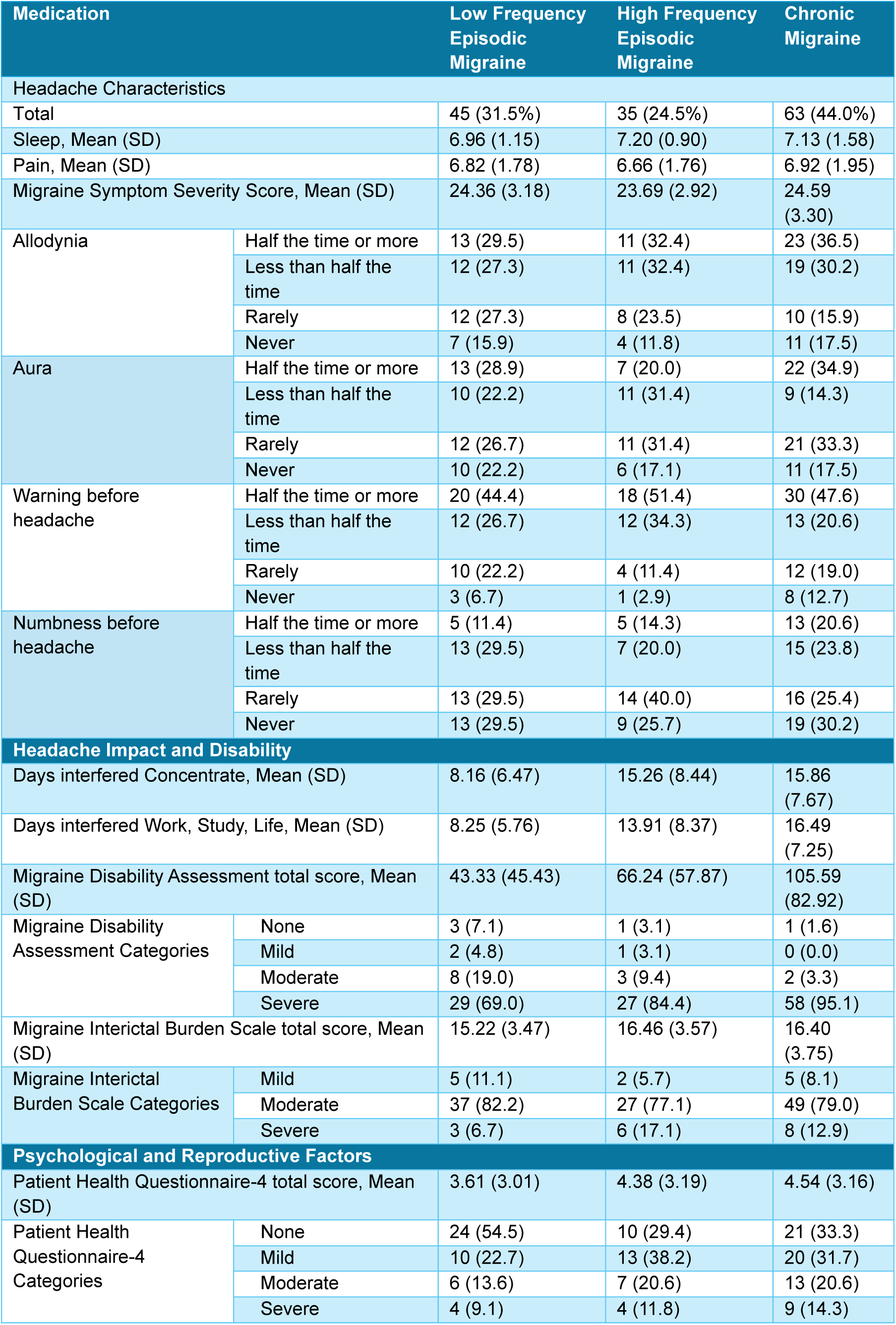
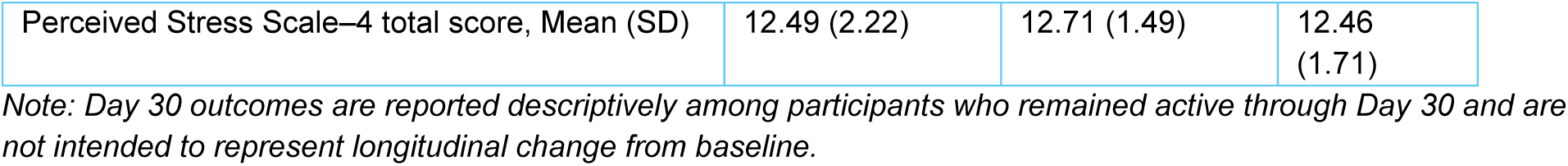
Patient reported outcomes in day 30 stratified by migraine type.

| Medication |  | Low Frequency Episodic Migraine | High Frequency Episodic Migraine | Chronic Migraine |
| --- | --- | --- | --- | --- |
| Headache Characteristics |  |  |  |  |
| Total |  | 45 (31.5%) | 35 (24.5%) | 63 (44.0%) |
| Sleep, Mean (SD) |  | 6.96 (1.15) | 7.20 (0.90) | 7.13 (1.58) |
| Pain, Mean (SD) |  | 6.82 (1.78) | 6.66 (1.76) | 6.92 (1.95) |
| Migraine Symptom Severity Score, Mean (SD) |  | 24.36 (3.18) | 23.69 (2.92) | 24.59 (3.30) |
| Allodynia | Half the time or more | 13 (29.5) | 11 (32.4) | 23 (36.5) |
|  | Less than half the time | 12 (27.3) | 11 (32.4) | 19 (30.2) |
|  | Rarely | 12 (27.3) | 8 (23.5) | 10 (15.9) |
|  | Never | 7 (15.9) | 4 (11.8) | 11 (17.5) |
| Aura | Half the time or more | 13 (28.9) | 7 (20.0) | 22 (34.9) |
|  | Less than half the time | 10 (22.2) | 11 (31.4) | 9 (14.3) |
|  | Rarely | 12 (26.7) | 11 (31.4) | 21 (33.3) |
|  | Never | 10 (22.2) | 6 (17.1) | 11 (17.5) |
| Warning before headache | Half the time or more | 20 (44.4) | 18 (51.4) | 30 (47.6) |
|  | Less than half the time | 12 (26.7) | 12 (34.3) | 13 (20.6) |
|  | Rarely | 10 (22.2) | 4 (11.4) | 12 (19.0) |
|  | Never | 3 (6.7) | 1 (2.9) | 8 (12.7) |
| Numbness before headache | Half the time or more | 5 (11.4) | 5 (14.3) | 13 (20.6) |
|  | Less than half the time | 13 (29.5) | 7 (20.0) | 15 (23.8) |
|  | Rarely | 13 (29.5) | 14 (40.0) | 16 (25.4) |
|  | Never | 13 (29.5) | 9 (25.7) | 19 (30.2) |
| Headache Impact and Disability |  |  |  |  |
| Days interfered Concentrate, Mean (SD) |  | 8.16 (6.47) | 15.26 (8.44) | 15.86 (7.67) |
| Days interfered Work, Study, Life, Mean (SD) |  | 8.25 (5.76) | 13.91 (8.37) | 16.49 (7.25) |
| Migraine Disability Assessment total score, Mean (SD) |  | 43.33 (45.43) | 66.24 (57.87) | 105.59 (82.92) |
| Migraine Disability Assessment Categories | None | 3 (7.1) | 1 (3.1) | 1 (1.6) |
|  | Mild | 2 (4.8) | 1 (3.1) | 0 (0.0) |
|  | Moderate | 8 (19.0) | 3 (9.4) | 2 (3.3) |
|  | Severe | 29 (69.0) | 27 (84.4) | 58 (95.1) |
| Migraine Interictal Burden Scale total score, Mean (SD) |  | 15.22 (3.47) | 16.46 (3.57) | 16.40 (3.75) |
| Migraine Interictal Burden Scale Categories | Mild | 5 (11.1) | 2 (5.7) | 5 (8.1) |
|  | Moderate | 37 (82.2) | 27 (77.1) | 49 (79.0) |
|  | Severe | 3 (6.7) | 6 (17.1) | 8 (12.9) |
| Psychological and Reproductive Factors |  |  |  |  |
| Patient Health Questionnaire-4 total score, Mean (SD) |  | 3.61 (3.01) | 4.38 (3.19) | 4.54 (3.16) |
| Patient Health Questionnaire-4 Categories | None | 24 (54.5) | 10 (29.4) | 21 (33.3) |
|  | Mild | 10 (22.7) | 13 (38.2) | 20 (31.7) |
|  | Moderate | 6 (13.6) | 7 (20.6) | 13 (20.6) |
|  | Severe | 4 (9.1) | 4 (11.8) | 9 (14.3) |

|  |  |  |  |
| --- | --- | --- | --- |
| Perceived Stress Scale—4 total score, Mean (SD) | 12.49 (2.22) | 12.71 (1.49) | 12.46<br>(1.71) |

*Note: Day 30 outcomes are reported descriptively among participants who remained active through Day 30 and are not intended to represent longitudinal change from baseline.*

**Supplementary Table 7.** Item-level completeness of EMA questionnaire responses, showing the proportion of missing responses for each item across all participants and study days.

| Feature | Skipped<br>(n) | Skipped<br>(%) |
| --- | --- | --- |
| Right now, do you have headache? | 28 | 0.0076 |
| What is the level of your headache pain intensity on a scale of 0-10? | 13 | 0.0114 |
| Is the pain worse on just one side? | 9 | 0.0079 |
| Is the pain pounding, pulsating or throbbing? | 10 | 0.0088 |
| Is the pain made worse by routine activities such as walking or climbing stairs? | 11 | 0.0097 |
| Do you feel nauseated? | 6 | 0.0053 |
| Does light bother you? | 11 | 0.0097 |
| Does sound bother you? | 12 | 0.0105 |
| Since last assessment, did you have headache? | 75 | 0.0203 |
| What was your average level of headache pain today on a scale of 0-10? | 13 | 0.0087 |
| How many days ago was your last migraine? | 4 | 0.0019 |
| What time did your headache start? | 52 | 0.0583 |
| How long did your headache last? | 7 | 0.0047 |
| How much did pain interfere with your daily activities? | 10 | 0.0067 |
| How much pain interfered with your ability to concentrate? | 12 | 0.008 |
| Did you feel that you have a brain fog and are not as sharp as usual today? | 42 | 0.0114 |
| Did you forget any tasks today? | 45 | 0.0122 |
| How much did forgetting tasks bother you? | 3 | 0.0037 |
| Have you been feeling moody today? | 38 | 0.0103 |
| How do you feel today? | 65 | 0.0176 |
| How many hours did you sleep last night? | 94 | 0.0255 |
| How was the quality of your sleep last night? | 49 | 0.0133 |
| Did you take any medications for migraine today? | 50 | 0.0136 |
| Please list any medications you have taken today for a headache/migraine | 53 | 0.0448 |
| Did you take a nap today? | 54 | 0.0146 |
| How long was your nap today? (In minutes) | 11 | 0.0077 |
| Overall, how enjoyable do you expect tomorrow will be? | 38 | 0.0103 |
| Overall, how stressful do you expect tomorrow will be? | 42 | 0.0114 |
| Did you take a sleep aid (such as Benadryl, melatonin, etc.) last night before going to sleep? | 42 | 0.0114 |

**Supplementary Table 8.**
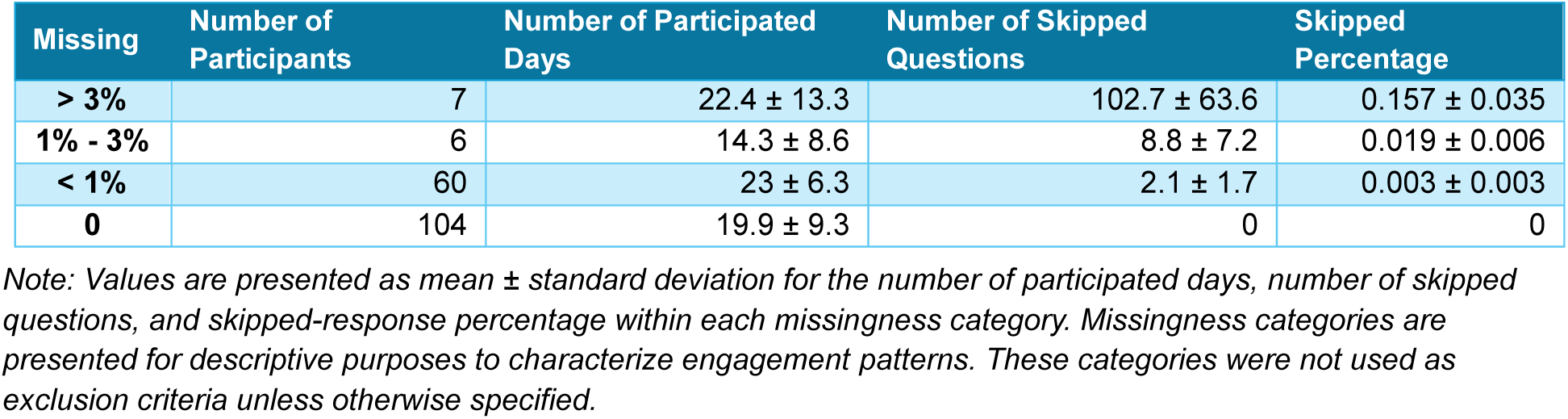
Distribution of participant compliance levels based on the proportion of missing responses.

| Missing | Number of<br>Participants | Number of Participated<br>Days | Number of Skipped<br>Questions | Skipped<br>Percentage |
| --- | --- | --- | --- | --- |
| > 3% | 7 | 22.4 ± 13.3 | 102.7 ± 63.6 | 0.157 ± 0.035 |
| 1% - 3% | 6 | 14.3 ± 8.6 | 8.8 ± 7.2 | 0.019 ± 0.006 |
| < 1% | 60 | 23 ± 6.3 | 2.1 ± 1.7 | 0.003 ± 0.003 |
| 0 | 104 | 19.9 ± 9.3 | 0 | 0 |

*Note: Values are presented as mean ± standard deviation for the number of participated days, number of skipped questions, and skipped-response percentage within each missingness category. Missingness categories are presented for descriptive purposes to characterize engagement patterns. These categories were not used as exclusion criteria unless otherwise specified.*

